## Supplemental Tables 1-6 for "Associations of Perceived Neighborhood Factors and Alzheimer’s Disease Polygenic Score with Cognition: Evidence from the Health and Retirement Study"

**Supplemental Table 1.** Characteristics of Included and Excluded Participants with Neighborhood disadvantage index in the Health and Retirement Study (2008-2010 Waves).

| Main Variables | Overall | Cognitive Impairment | | | CIND | | | Dementia | | |
|---|---|---|---|---|---|---|---|---|---|---|
| | **(N=14,562)**[a] | **Excluded sample (N=7,736)** [a] | **Included sample (N=6,826)** [a] | **p-value**[b] | **Excluded sample (N=7,816)** [a] | **Included sample (N=6,746)** [a] | **p-value**[b] | **Excluded sample (N=6,802)** [a] | **Included sample (N=7,760)** [a] | **p-value**[b] |
| **CIND & Dementia** | | | | **<0.001** | | | **<0.001** | | | **<0.001** |
| Yes | 5,453 (41%) | 3,381 (53%) | 2,072 (30%) | | 3,461 (54%) | 1,992 (30%) | | 2,582 (48%) | 2,871 (37%) | |
| No | 7,700 (59%) | 2,946 (47%) | 4,754 (70%) | | 2,946 (46%) | 4,754 (70%) | | 2,811 (52%) | 4,889 (63%) | |
| Missing | 1,409 | 1,409 | | | 1,409 | | | 1,409 | | |
| **CIND** | | | | | | | **<0.001** | | | **<0.001** |
| Yes | 5,010 (38%) | 3,018 (48%) | 1,992 (29%) | **<0.001** | 3,018 (47%) | 1,992 (30%) | | 2,332 (43%) | 2,678 (35%) | |
| No | 8,143 (62%) | 3,309 (52%) | 4,834 (71%) | | 3,389 (53%) | 4,754 (70%) | | 3,061 (57%) | 5,082 (65%) | |
| Missing | 1,409 | 1,409 | | | 1,409 | | | 1,409 | | |
| **Dementia** | | | | **<0.001** | | | **<0.001** | | | **<0.001** |
| Yes | 1,414 (11%) | 1,125 (18%) | 289 (4.2%) | | 1,205 (19%) | 209 (3.1%) | | 796 (15%) | 618 (8.0%) | |
| No | 11,739 (89%) | 5,202 (82%) | 6,537 (96%) | | 5,202 (81%) | 6,537 (97%) | | 4,597 (85%) | 7,142 (92%) | |
| Missing | 1,409 | 1,409 | | | 1,409 | | | 1,409 | | |
| **Neighborhood disadvantage index** | 0.00 (1.00) | 0.11 (1.07) | -0.13 (0.90) | **<0.001** | 0.11 (1.07) | -0.13 (0.90) | **<0.001** | 0.11 (1.07) | -0.10 (0.93) | **<0.001** |
| **Neighborhood disadvantage index (Binary)** | | | | **<0.001** | | | **<0.001** | | | **<0.001** |
| The least disadvantaged neighborhoods (<=0) | 8,917 (61%) | 4,380 (57%) | 4,537 (66%) | | 4,429 (57%) | 4,488 (67%) | | 3,847 (57%) | 5,070 (65%) | |
| The most disadvantaged neighborhoods (>0) | 5,645 (39%) | 3,356 (43%) | 2,289 (34%) | | 3,387 (43%) | 2,258 (33%) | | 2,955 (43%) | 2,690 (35%) | |
| **Baseline wave** | | | | **<0.001** | | | **<0.001** | | | **<0.001** |
| Wave 1 (2008) | 6,714 (46%) | 3,706 (48%) | 3,008 (44%) | | 3,743 (48%) | 2,971 (44%) | | 3,278 (48%) | 3,436 (44%) | |
| Wave 2 (2010) | 7,848 (54%) | 4,030 (52%) | 3,818 (56%) | | 4,073 (52%) | 3,775 (56%) | | 3,524 (52%) | 4,324 (56%) | |
| **Polygenic score for AD** | | | | | | | | | | |
| European ancestry | -0.02 (1.00) | 0.14 (1.04) | -0.08 (0.98) | - | 0.14 (1.04) | -0.09 (0.98) | - | 0.20 (1.05) | -0.08 (0.98) | - |
| Missing | 5,685 | 4,982 | 703 | | 4,990 | 695 | | 4,714 | 971 | |
| African ancestry | 0.02 (0.99) | -0.01 (1.03) | 0.06 (0.93) | - | -0.01 (1.03) | 0.06 (0.93) | - | -0.04 (1.11) | 0.06 (0.91) | - |
| Missing | 13,031 | 6908 | 6123 | | 6,980 | 6,051 | | 6,242 | 6,789 | |
| **Polygenic score for AD (Binary)** | | | | | | | | | | |
| European ancestry | | | | - | | | - | | | - |
| Below 75% | 6,657 (75%) | 1,888 (69%) | 4,769 (78%) | | 1,938 (69%) | 4,719 (78%) | | 1,390 (67%) | 5,267 (78%) | |
| Above 75% | 2,220 (25%) | 866 (31%) | 1,354 (22%) | | 888 (31%) | 1,332 (22%) | | 698 (33%) | 1,522 (22%) | |
| Missing | 5,685 | 4,982 | 703 | | 4,990 | 695 | | 4,714 | 971 | |
| African ancestry | | | | - | | | - | | | - |
| Below 75% | 1,148 (75%) | 614 (74%) | 534 (76%) | | 617 (74%) | 531 (76%) | | 407 (73%) | 741 (76%) | |
| Above 75% | 383 (25%) | 214 (26%) | 169 (24%) | | 219 (26%) | 164 (24%) | | 153 (27%) | 230 (24%) | |
| Missing | 13,031 | 6,908 | 6,123 | | 6,980 | 6,051 | | 6,242 | 6,789 | |
| **APOE E4 status** | | | | 0.706 | | | 0.623 | | | 0.185 |
| Any copies of e4 | 3,089 (27%) | 1,282 (27%) | 1,807 (26%) | | 1,306 (27%) | 1,783 (26%) | | 995 (26%) | 2,094 (27%) | |
| No copies of e4 | 8,523 (73%) | 3,504 (73%) | 5,019 (74%) | | 3,560 (73%) | 4,963 (74%) | | 2,857 (74%) | 5,666 (73%) | |
| Missing | 2,950 | 2,950 | | | 2,950 | | | 2,950 | | |
| **Sex** | | | | **<0.001** | | | **0.018** | | | **<0.001** |
| Male | 6,011 (41%) | 3,337 (43%) | 2,674 (39%) | | 3,370 (43%) | 2,641 (39%) | | 2,918 (43%) | 3,093 (40%) | |
| Female | 8,551 (59%) | 4,399 (57%) | 4,152 (61%) | | 4,446 (57%) | 4,105 (61%) | | 3,884 (57%) | 4,667 (60%) | |
| **Age** | 67.57 (10.92) | 68.42 (11.56) | 66.61 (10.06) | **<0.001** | 68.46 (11.56) | 66.54 (10.04) | **<0.001** | 67.89 (11.61) | 67.29 (10.28) | **0.001** |
| **Education Level** | | | | **<0.001** | | | **<0.001** | | | **<0.001** |
| > High School / GED | 4,128 (28%) | 1,766 (23%) | 2,362 (35%) | | 1,792 (23%) | 2,336 (35%) | | 1,639 (24%) | 2,489 (32%) | |
| High School / GED | 7,895 (54%) | 3,974 (51%) | 3,921 (57%) | | 4,019 (51%) | 3,876 (57%) | | 3,470 (51%) | 4,425 (57%) | |
| < High School / GED | 2,536 (17%) | 1,993 (26%) | 543 (8.0%) | | 2,002 (26%) | 534 (7.9%) | | 1,690 (25%) | 846 (11%) | |
| Missing | 3 | 3 | | | 3 | | | 3 | | |
| **Poverty Status** | | | | **<0.001** | | | **<0.001** | | | **<0.001** |
| Above Poverty threshold | 13,179 (91%) | 6,661 (87%) | 6,518 (95%) | | 6,738 (87%) | 6,441 (95%) | | 5,859 (88%) | 7,320 (94%) | |
| Below Poverty threshold | 1,270 (8.8%) | 962 (13%) | 308 (4.5%) | | 965 (13%) | 305 (4.5%) | | 830 (12%) | 440 (5.7%) | |
| Missing | 113 | 113 | | | 113 | | | 113 | | |
| **Social Ladder** | 6.41 (1.76) | 6.26 (1.85) | 6.55 (1.66) | **<0.001** | 6.27 (1.85) | 6.55 (1.66) | **<0.001** | 6.27 (1.85) | 6.51 (1.68) | **<0.001** |
| Missing | 1,367 | 1,367 | | | 1,367 | | | 1,367 | | |
| **Sensitivity Variables** | **Overall** | **Excluded sample** | **Included sample** | **p-value** | **Excluded sample** | **Included sample** | **p-value** | **Excluded sample** | **Included sample** | **p-value** |
| **Smoking status** | | | | **<0.001** | | | **<0.001** | | | **<0.001** |
| Current Smoker | 1,968 (14%) | 1,149 (15%) | 819 (12%) | | 1,155 (15%) | 813 (12%) | | 1,022 (15%) | 946 (12%) | |
| Former Smoker | 6,188 (43%) | 3,271 (43%) | 2,917 (43%) | | 3,311 (43%) | 2,877 (43%) | | 2,830 (42%) | 3,358 (44%) | |
| Never Smoke | 6,307 (44%) | 3,257 (42%) | 3,050 (45%) | | 3,289 (42%) | 3,018 (45%) | | 2,896 (43%) | 3,411 (44%) | |
| Missing | 99 | 59 | 40 | | 61 | 38 | | 54 | 45 | |
| **Drinking (# drinks/day when drinks)** | 0.76 (1.47) | 0.72 (1.55) | 0.81 (1.37) | **<0.001** | 0.72 (1.55) | 0.81 (1.37) | **<0.001** | 0.73 (1.56) | 0.78 (1.38) | **0.026** |
| Missing | 14 | 10 | 4 | | 10 | 4 | | 9 | 5 | |
| **BMI** | 28.43 (6.02) | 28.22 (6.02) | 28.67 (6.00) | **<0.001** | 28.21 (6.02) | 28.68 (6.00) | **<0.001** | 28.23 (5.99) | 28.60 (6.04) | **<0.001** |
| Missing | 178 | 127 | 51 | | 128 | 50 | | 117 | 61 | |
| **Ever have Diabetes** | | | | **<0.001** | | | | | | **<0.001** |
| Yes | 2,979 (20%) | 1,850 (24%) | 1,129 (17%) | | 1,860 (24%) | 1,119 (17%) | | 1,610 (24%) | 1,369 (18%) | |
| No | 11,583 (80%) | 5,886 (76%) | 5,697 (83%) | | 5,956 (76%) | 5,627 (83%) | | 5,192 (76%) | 6,391 (82%) | |
| **Chronic Condition** | | | | **<0.001** | | | **<0.001** | | | **<0.001** |
| None | 2,149 (15%) | 1,093 (14%) | 1,056 (15%) | | 1,104 (14%) | 1,045 (15%) | | 993 (15%) | 1,156 (15%) | |
| 2-Jan | 7,251 (50%) | 3,621 (47%) | 3,630 (53%) | | 3,656 (47%) | 3,595 (53%) | | 3,218 (47%) | 4,033 (52%) | |
| >= 3 | 5,162 (35%) | 3,022 (39%) | 2,140 (31%) | | 3,056 (39%) | 2,106 (31%) | | 2,591 (38%) | 2,571 (33%) | |
| **Depression** | 1.38 (1.93) | 1.66 (2.09) | 1.08 (1.71) | **<0.001** | 1.66 (2.08) | 1.08 (1.71) | **<0.001** | 1.38 (1.93) | 1.66 (2.09) | **<0.001** |
| Missing | 415 | 415 | | | 415 | | | 415 | | |

[a]n (%); Mean (SD)

[b]Pearson's Chi-squared test; Welch Two Sample t-test

**Supplemental Table 2**: Sample distribution by genetic ancestry in the Health and Retirement Study, wave 2008-2010.

| Main Variables | Cognitive Impairment | | | CIND | | | Dementia | | |
|---|---|---|---|---|---|---|---|---|---|
| | **Study sample (N=6,826)** [a] | **African ancestry (N=703)** [a] | **European ancestry (N=6,123)** [a] | **Study sample (N=6,746)** [a] | **African ancestry (N=695)** [a] | **European ancestry (N=6,051)** [a] | **Study sample (N=7,760)** [a] | **African ancestry (N=971)** [a] | **European ancestry (N=6,789)** [a] |
| **Neighborhood disadvantage index** | -0.13 (0.90) | 0.47 (1.04) | -0.20 (0.85) | -0.13 (0.90) | 0.47 (1.04) | -0.20 (0.85) | -0.10 (0.93) | 0.49 (1.06) | -0.18 (0.87) |
| **Neighborhood disadvantage index (Binary)** | | | | | | | | | |
| The least disadvantaged neighborhoods (<=0) | 4,537 (66%) | 274 (39%) | 4,263 (70%) | 4,488 (67%) | 272 (39%) | 4,216 (70%) | 5,070 (65%) | 372 (38%) | 4,698 (69%) |
| The most disadvantaged neighborhoods (>0) | 2,289 (34%) | 429 (61%) | 1,860 (30%) | 2,258 (33%) | 423 (61%) | 1,835 (30%) | 2,690 (35%) | 599 (62%) | 2,091 (31%) |
| **PGS-AD** | | | | | | | | | |
| European ancestry | -0.08 (0.98) | - | -0.08 (0.98) | -0.09 (0.98) | - | -0.09 (0.98) | -0.08 (0.98) | - | -0.08 (0.98) |
| African ancestry | 0.06 (0.93) | 0.06 (0.93) | - | 0.06 (0.93) | 0.06 (0.93) | - | 0.06 (0.91) | 0.06 (0.91) | - |
| **PGS-AD (Binary)** | | | | | | | | | |
| European ancestry | | | | | | | | | |
| Below 75% | 4,769 (78%) | - | 4,769 (78%) | 4,719 (78%) | - | 4,719 (78%) | 5,267 (78%) | - | 5,267 (78%) |
| Above 75% | 1,354 (22%) | - | 1,354 (22%) | 1,332 (22%) | - | 1,332 (22%) | 1,522 (22%) | - | 1,522 (22%) |
| African ancestry | | | | | | | | | |
| Below 75% | 534 (76%) | 534 (76%) | - | 531 (76%) | 531 (76%) | - | 741 (76%) | 741 (76%) | - |
| Above 75% | 169 (24%) | 169 (24%) | - | 164 (24%) | 164 (24%) | - | 230 (24%) | 230 (24%) | - |
| **APOE E4 status** | | | | | | | | | |
| Any copies of e4 | 1,807 (26%) | 261 (37%) | 1,546 (25%) | 1,783 (26%) | 257 (37%) | 1,526 (25%) | 2,094 (27%) | 356 (37%) | 1,738 (26%) |
| No copies of e4 | 5,019 (74%) | 442 (63%) | 4,577 (75%) | 4,963 (74%) | 438 (63%) | 4,525 (75%) | 5,666 (73%) | 615 (63%) | 5,051 (74%) |
| **Sex** | | | | | | | | | |
| Female | 4,152 (61%) | 491 (70%) | 3,661 (60%) | 4,105 (61%) | 485 (70%) | 3,620 (60%) | 4,667 (60%) | 653 (67%) | 4,014 (59%) |
| Male | 2,674 (39%) | 212 (30%) | 2,462 (40%) | 2,641 (39%) | 210 (30%) | 2,431 (40%) | 3,093 (40%) | 318 (33%) | 2,775 (41%) |
| **Age** | 66.61 (10.06) | 62.06 (8.84) | 67.14 (10.06) | 66.54 (10.04) | 62.08 (8.86) | 67.05 (10.04) | 67.29 (10.28) | 63.67 (9.56) | 67.81 (10.27) |
| **Education Level** | | | | | | | | | |
| > High School / GED | 2,362 (35%) | 195 (28%) | 2,167 (35%) | 2,336 (35%) | 192 (28%) | 2,144 (35%) | 2,489 (32%) | 210 (22%) | 2,279 (34%) |
| High School / GED | 3,921 (57%) | 393 (56%) | 3,528 (58%) | 3,876 (57%) | 388 (56%) | 3,488 (58%) | 4,425 (57%) | 528 (54%) | 3,897 (57%) |
| < High School / GED | 543 (8.0%) | 115 (16%) | 428 (7.0%) | 534 (7.9%) | 115 (17%) | 419 (6.9%) | 846 (11%) | 233 (24%) | 613 (9.0%) |
| **Poverty Status** | | | | | | | | | |
| Above Poverty threshold | 6,518 (95%) | 594 (84%) | 5,924 (97%) | 6,441 (95%) | 587 (84%) | 5,854 (97%) | 7,320 (94%) | 783 (81%) | 6,537 (96%) |
| Below Poverty threshold | 308 (4.5%) | 109 (16%) | 199 (3.3%) | 305 (4.5%) | 108 (16%) | 197 (3.3%) | 440 (5.7%) | 188 (19%) | 252 (3.7%) |
| **Baseline wave** | | | | | | | | | |
| Wave 1 (2008) | 3,008 (44%) | 273 (39%) | 2,735 (45%) | 2,971 (44%) | 271 (39%) | 2,700 (45%) | 3,436 (44%) | 391 (40%) | 3,045 (45%) |
| Wave 2 (2010) | 3,818 (56%) | 430 (61%) | 3,388 (55%) | 3,775 (56%) | 424 (61%) | 3,351 (55%) | 4,324 (56%) | 580 (60%) | 3,744 (55%) |
| **Social Ladder** | -0.08 (0.94) | 0.24 (1.02) | -0.11 (0.93) | 6.55 (1.66) | 5.99 (1.80) | 6.61 (1.63) | 6.51 (1.68) | 5.95 (1.83) | 6.59 (1.65) |
| **Sensitivity Variables** | **Sensitivity analytic sample** | **African ancestry** | **European ancestry** | **Sensitivity analytic sample** | **African ancestry** | **European ancestry** | **Sensitivity analytic sample** | **African ancestry** | **European ancestry** |
| **Smoking status** | | | | | | | | | |
| Current Smoker | 819 (12%) | 139 (20%) | 680 (11%) | 813 (12%) | 138 (20%) | 675 (11%) | 946 (12%) | 186 (19%) | 760 (11%) |
| Former Smoker | 2,917 (43%) | 266 (38%) | 2,651 (44%) | 2,877 (43%) | 262 (38%) | 2,615 (43%) | 3,358 (44%) | 386 (40%) | 2,972 (44%) |
| Never Smoke | 3,050 (45%) | 296 (42%) | 2,754 (45%) | 3,018 (45%) | 293 (42%) | 2,725 (45%) | 3,411 (44%) | 395 (41%) | 3,016 (45%) |
| **BMI** | 28.67 (6.00) | 31.13 (7.28) | 28.38 (5.77) | 28.68 (6.00) | 31.14 (7.28) | 28.40 (5.77) | 28.60 (6.04) | 30.86 (7.14) | 28.28 (5.79) |
| **Drinking (# drinks/day when drinks)** | 0.81 (1.37) | 0.66 (1.31) | 0.82 (1.37) | 0.81 (1.37) | 0.66 (1.32) | 0.83 (1.38) | 0.78 (1.38) | 0.64 (1.33) | 0.81 (1.39) |
| **Ever have Diabetes** | | | | | | | | | |
| Yes | 1,129 (17%) | 168 (24%) | 961 (16%) | 1,119 (17%) | 166 (24%) | 953 (16%) | 1,369 (18%) | 264 (27%) | 1,105 (16%) |
| No | 5,697 (83%) | 535 (76%) | 5,162 (84%) | 5,627 (83%) | 529 (76%) | 5,098 (84%) | 6,391 (82%) | 707 (73%) | 5,684 (84%) |
| **Chronic Condition** | | | | | | | | | |
| None | 1,056 (15%) | 104 (15%) | 952 (16%) | 1,045 (15%) | 103 (15%) | 942 (16%) | 1,156 (15%) | 140 (14%) | 1,016 (15%) |
| 1 - 2 | 3,630 (53%) | 378 (54%) | 3,252 (53%) | 3,595 (53%) | 373 (54%) | 3,222 (53%) | 4,033 (52%) | 479 (49%) | 3,554 (52%) |
| >= 3 | 2,140 (31%) | 221 (31%) | 1,919 (31%) | 2,106 (31%) | 219 (32%) | 1,887 (31%) | 2,571 (33%) | 352 (36%) | 2,219 (33%) |
| **Depression** | 1.08 (1.71) | 1.47 (1.90) | 1.03 (1.68) | 1.08 (1.71) | 1.47 (1.90) | 1.03 (1.68) | 1.15 (1.76) | 1.62 (2.00) | 1.09 (1.72) |

[a]n (%); Mean (SD)

**Supplemental Table 3:** Hazard Ratios from survival analysis with binary Neighborhood disadvantage index stratified by Ancestry, estimates present the association for the most Disadvantage Neighborhoods with incident cognitive impairment (CIND and dementia), CIND and dementia, relative to normal cognition and non-dementia respectively in the US Health and Retirement Study (2008-2010 Waves).

| | **Cognitive Impairment vs. Normal Cognition, European Ancestry (n=6,123)** | | | | | | | | |
|---|---|---|---|---|---|---|---|---|---|
| | **Model 1** | | | **Model 2** | | | **Model 3** | | |
| | **HR** | **95% CI** | **p-value** | **HR** | **95% CI** | **p-value** | **HR** | **95% CI** | **p-value** |
| **Neighborhood disadvantage index** | | | | | | | | | |
| The least disadvantaged neighborhoods (<=0) | Ref | Ref | Ref | Ref | Ref | Ref | Ref | Ref | Ref |
| The most disadvantaged neighborhoods (>0) | 1.19 | 1.08, 1.32 | **<0.001** | 1.20 | 1.08, 1.33 | **<0.001** | 1.21 | 1.08, 1.36 | **0.001** |
| **Age** | 1.08 | 1.07, 1.08 | **<0.001** | 1.08 | 1.07, 1.08 | **<0.001** | 1.08 | 1.07, 1.08 | **<0.001** |
| **Sex** | | | | | | | | | |
| Female | Ref | Ref | Ref | Ref | Ref | Ref | Ref | Ref | Ref |
| Male | 1.31 | 1.19, 1.44 | **<0.001** | 1.30 | 1.18, 1.43 | **<0.001** | 1.30 | 1.18, 1.43 | **<0.001** |
| **Education** | | | | | | | | | |
| Above High School/GED | Ref | Ref | Ref | Ref | Ref | Ref | Ref | Ref | Ref |
| High School/GED | 1.62 | 1.44, 1.82 | **<0.001** | 1.62 | 1.44, 1.82 | **<0.001** | 1.62 | 1.44, 1.82 | **<0.001** |
| Less than High School/GED | 3.04 | 2.57, 3.60 | **<0.001** | 3.04 | 2.57, 3.59 | **<0.001** | 3.04 | 2.57, 3.59 | **<0.001** |
| **Poverty Status (Below)** | | | | | | | | | |
| Above Poverty threshold | Ref | Ref | Ref | Ref | Ref | Ref | Ref | Ref | Ref |
| Below Poverty threshold | 1.28 | 0.99, 1.65 | 0.056 | 1.27 | 0.99, 1.64 | 0.063 | 1.27 | 0.99, 1.64 | 0.063 |
| **APOE E4 status** | | | | | | | | | |
| No copies of e4 | Ref | Ref | Ref | Ref | Ref | Ref | Ref | Ref | Ref |
| Any copies of e4 | 1.43 | 1.29, 1.59 | **<0.001** | 1.43 | 1.29, 1.59 | **<0.001** | 1.43 | 1.28, 1.58 | **<0.001** |
| **Social Ladder** | 0.93 | 0.90, 0.95 | **<0.001** | 0.93 | 0.90, 0.95 | **<0.001** | 0.93 | 0.90, 0.95 | **<0.001** |
| **Baseline wave** | | | | | | | | | |
| Wave 1 (2008) | Ref | Ref | Ref | Ref | Ref | Ref | Ref | Ref | Ref |
| Wave 2 (2010) | 0.86 | 0.78, 0.94 | **0.001** | 0.86 | 0.78, 0.95 | **0.002** | 0.86 | 0.78, 0.94 | **0.002** |
| **PGS-AD** | | | | | | | | | |
| Below 75% | - | - | - | Ref | Ref | Ref | Ref | Ref | Ref |
| Above 75% | - | - | - | 1.12 | 1.01, 1.25 | **0.040** | 1.14 | 0.99, 1.30 | 0.061 |
| **Neighborhood* PGS-AD** | - | - | - | - | - | - | 0.96 | 0.76, 1.22 | 0.700 |
| RERI: The most disadvantaged neighborhoods*PGS-AD Above 75% | - | - | - | - | - | - | -0.02 | -0.31, 0.27 | |

| | **CIND vs. Normal Cognition, European Ancestry (n=6,051)** | | | | | | | | |
|---|---|---|---|---|---|---|---|---|---|
| | **Model 1** | | | **Model 2** | | | **Model 3** | | |
| | **HR** | **95% CI** | **p-value** | **HR** | **95% CI** | **p-value** | **HR** | **95% CI** | **p-value** |
| **Neighborhood disadvantage index** | | | | | | | | | |
| The least disadvantaged neighborhoods (<=0) | Ref | Ref | Ref | Ref | Ref | Ref | Ref | Ref | Ref |
| The most disadvantaged neighborhoods (>0) | 1.18 | 1.06, 1.31 | **0.002** | 1.19 | 1.07, 1.31 | **0.001** | 1.20 | 1.06, 1.35 | **0.003** |
| **Age** | 1.08 | 1.07, 1.08 | **<0.001** | 1.08 | 1.07, 1.08 | **<0.001** | 1.08 | 1.07, 1.08 | **<0.001** |
| **Sex** | | | | | | | | | |
| Female | Ref | Ref | Ref | Ref | Ref | Ref | Ref | Ref | Ref |
| Male | 1.32 | 1.19, 1.45 | **<0.001** | 1.31 | 1.19, 1.45 | **<0.001** | 1.31 | 1.19, 1.45 | **<0.001** |
| **Education** | | | | | | | | | |
| Above High School/GED | Ref | Ref | Ref | Ref | Ref | Ref | Ref | Ref | Ref |
| High School/GED | 1.65 | 1.46, 1.86 | **<0.001** | 1.65 | 1.47, 1.86 | **<0.001** | 1.65 | 1.47, 1.86 | **<0.001** |
| Less than High School/GED | 3.09 | 2.61, 3.67 | **<0.001** | 3.10 | 2.61, 3.67 | **<0.001** | 3.09 | 2.61, 3.67 | **<0.001** |
| **Poverty Status (Below)** | | | | | | | | | |
| Above Poverty threshold | Ref | Ref | Ref | Ref | Ref | Ref | Ref | Ref | Ref |
| Below Poverty threshold | 1.28 | 0.99, 1.66 | 0.060 | 1.27 | 0.98, 1.65 | 0.069 | 1.27 | 0.98, 1.65 | 0.069 |
| **APOE E4 status** | | | | | | | | | |
| No copies of e4 | Ref | Ref | Ref | Ref | Ref | Ref | Ref | Ref | Ref |
| Any copies of e4 | 1.44 | 1.29, 1.60 | **<0.001** | 1.43 | 1.28, 1.59 | **<0.001** | 1.43 | 1.28, 1.59 | **<0.001** |
| **Social Ladder** | 0.92 | 0.90, 0.95 | **<0.001** | 0.92 | 0.89, 0.95 | **<0.001** | 0.92 | 0.89, 0.95 | **<0.001** |
| **Baseline wave** | | | | | | | | | |
| Wave 1 (2008) | Ref | Ref | Ref | Ref | Ref | Ref | Ref | Ref | Ref |
| Wave 2 (2010) | 0.86 | 0.78, 0.95 | **0.002** | 0.86 | 0.78, 0.95 | **0.002** | 0.86 | 0.78, 0.95 | **0.002** |
| **PGS-AD** | | | | | | | | | |
| Below 75% | - | - | - | Ref | Ref | Ref | Ref | Ref | Ref |
| Above 75% | - | - | - | 1.12 | 1.00, 1.25 | 0.055 | 1.13 | 0.99, 1.30 | 0.078 |
| **Neighborhood* PGS-AD** | - | - | - | - | - | - | 0.96 | 0.76, 1.22 | 0.700 |
| RERI: The most disadvantaged neighborhoods*PGS-AD Above 75% | - | - | - | - | - | - | -0.03 | -0.32, 0.27 | |

| | **Dementia vs. Non-dementia, European Ancestry (n=6,789)** | | | | | | | | |
|---|---|---|---|---|---|---|---|---|---|
| | **Model 1** | | | **Model 2** | | | **Model 3** | | |
| | **HR** | **95% CI** | **p-value** | **HR** | **95% CI** | **p-value** | **HR** | **95% CI** | **p-value** |
| **Neighborhood disadvantage index** | | | | | | | | | |
| The least disadvantaged neighborhoods (<=0) | Ref | Ref | Ref | Ref | Ref | Ref | Ref | Ref | Ref |
| The most disadvantaged neighborhoods (>0) | 1.34 | 1.10, 1.63 | **0.004** | 1.33 | 1.09, 1.62 | **0.005** | 1.34 | 1.07, 1.68 | **0.011** |
| **Age** | 1.12 | 1.10, 1.13 | **<0.001** | 1.12 | 1.10, 1.13 | **<0.001** | 1.12 | 1.10, 1.13 | **<0.001** |
| **Sex** | | | | | | | | | |
| Female | Ref | Ref | Ref | Ref | Ref | Ref | Ref | Ref | Ref |
| Male | 1.06 | 0.87, 1.28 | 0.600 | 1.06 | 0.88, 1.29 | 0.500 | 1.06 | 0.87, 1.29 | 0.500 |
| **Education** | | | | | | | | | |
| Above High School/GED | Ref | Ref | Ref | Ref | Ref | Ref | Ref | Ref | Ref |
| High School/GED | 1.59 | 1.23, 2.05 | **<0.001** | 1.61 | 1.24, 2.07 | **<0.001** | 1.61 | 1.24, 2.07 | **<0.001** |
| Less than High School/GED | 3.99 | 2.96, 5.37 | **<0.001** | 4.04 | 3.00, 5.44 | **<0.001** | 4.04 | 3.00, 5.44 | **<0.001** |
| **Poverty Status (Below)** | | | | | | | | | |
| Above Poverty threshold | Ref | Ref | Ref | Ref | Ref | Ref | Ref | Ref | Ref |
| Below Poverty threshold | 2.04 | 1.39, 2.99 | **<0.001** | 2.07 | 1.41, 3.04 | **<0.001** | 2.07 | 1.41, 3.04 | **<0.001** |
| **APOE E4 status** | | | | | | | | | |
| No copies of e4 | Ref | Ref | Ref | Ref | Ref | Ref | Ref | Ref | Ref |
| Any copies of e4 | 2.06 | 1.70, 2.50 | **<0.001** | 2.05 | 1.69, 2.49 | **<0.001** | 2.05 | 1.69, 2.49 | **<0.001** |
| **Social Ladder** | 0.98 | 0.93, 1.05 | 0.600 | 0.99 | 0.93, 1.05 | 0.700 | 0.99 | 0.93, 1.05 | 0.700 |
| **Baseline wave** | | | | | | | | | |
| Wave 1 (2008) | Ref | Ref | Ref | Ref | Ref | Ref | Ref | Ref | Ref |
| Wave 2 (2010) | 0.81 | 0.67, 0.97 | **0.023** | 0.8 | 0.67, 0.97 | **0.020** | 0.8 | 0.67, 0.97 | **0.020** |
| **PGS-AD** | | | | | | | | | |
| Below 75% | - | - | - | Ref | Ref | Ref | Ref | Ref | Ref |
| Above 75% | - | - | - | 1.19 | 0.96, 1.47 | 0.120 | 1.20 | 0.92, 1.57 | 0.200 |
| **Neighborhood* PGS-AD** | - | - | - | - | - | - | 0.96 | 0.62, 1.51 | 0.900 |
| RERI: The most disadvantaged neighborhoods*PGS-AD Above 75% | - | - | - | - | - | - | 0.01 | -0.61, 0.63 | |

| | **Cognitive Impairment vs. Normal Cognition, African Ancestry (n=703)** | | | | | | | | |
|---|---|---|---|---|---|---|---|---|---|
| | **Model 1** | | | **Model 2** | | | **Model 3** | | |
| | **HR** | **95% CI** | **p-value** | **HR** | **95% CI** | **p-value** | **HR** | **95% CI** | **p-value** |
| **Neighborhood disadvantage index** | | | | | | | | | |
| The least disadvantaged neighborhoods (<=0) | Ref | Ref | Ref | Ref | Ref | Ref | Ref | Ref | Ref |
| The most disadvantaged neighborhoods (>0) | 1.05 | 0.83, 1.33 | 0.700 | 1.02 | 0.81, 1.30 | 0.800 | 1.13 | 0.86, 1.50 | 0.4 |
| **Age** | 1.05 | 1.04, 1.07 | **<0.001** | 1.05 | 1.04, 1.07 | **<0.001** | 1.05 | 1.04, 1.07 | **<0.001** |
| **Sex** | | | | | | | | | |
| Female | Ref | Ref | Ref | Ref | Ref | Ref | Ref | Ref | Ref |
| Male | 1.19 | 0.93, 1.52 | 0.200 | 1.21 | 0.94, 1.54 | 0.130 | 1.21 | 0.94, 1.55 | 0.13 |
| **Education** | | | | | | | | | |
| Above High School/GED | Ref | Ref | Ref | Ref | Ref | Ref | Ref | Ref | Ref |
| High School/GED | 1.51 | 1.12, 2.05 | **0.008** | 1.51 | 1.11, 2.04 | **0.009** | 1.5 | 1.11, 2.04 | **0.009** |
| Less than High School/GED | 3.04 | 2.13, 4.33 | **<0.001** | 2.96 | 2.07, 4.23 | **<0.001** | 2.99 | 2.09, 4.27 | **<0.001** |
| **Poverty Status** | | | | | | | | | |
| Above Poverty threshold | Ref | Ref | Ref | Ref | Ref | Ref | Ref | Ref | Ref |
| Below Poverty threshold | 1.78 | 1.33, 2.36 | **<0.001** | 1.79 | 1.34, 2.39 | **<0.001** | 1.79 | 1.35, 2.39 | **<0.001** |
| **APOE E4 status** | | | | | | | | | |
| No copies of e4 | Ref | Ref | Ref | Ref | Ref | Ref | Ref | Ref | Ref |
| Any copies of e4 | 1.02 | 0.80, 1.29 | 0.900 | 1.02 | 0.80, 1.29 | >0.9 | 1.01 | 0.80, 1.28 | >0.9 |
| **Social Ladder** | 0.97 | 0.91, 1.03 | 0.300 | 0.96 | 0.89, 1.02 | 0.200 | 0.96 | 0.90, 1.02 | **0.2** |
| **Baseline wave** | | | | | | | | | |
| Wave 1 (2008) | Ref | Ref | Ref | Ref | Ref | Ref | Ref | Ref | Ref |
| Wave 2 (2010) | 1.19 | 0.93, 1.51 | 0.200 | 1.17 | 0.92, 1.49 | 0.200 | 1.16 | 0.91, 1.48 | 0.2 |
| **PGS-AD** | | | | | | | | | |
| Below 75% | - | - | - | Ref | Ref | Ref | Ref | Ref | Ref |
| Above 75% | - | - | - | 1.11 | 0.84, 1.48 | 0.500 | 1.39 | 0.91, 2.13 | 0.13 |
| **Neighborhood* PGS-AD** | - | - | - | - | - | - | 0.7 | 0.42, 1.17 | 0.2 |
| RERI: The most disadvantaged neighborhoods*PGS-AD Above 75% | - | - | - | - | - | - | -0.42 | -1.09, 0.25 | |

| | **CIND vs. Normal Cognition, African Ancestry (n=695)** | | | | | | | | |
|---|---|---|---|---|---|---|---|---|---|
| | **Model 1** | | | **Model 2** | | | **Model 3** | | |
| | **HR** | **95% CI** | **p-value** | **HR** | **95% CI** | **p-value** | **HR** | **95% CI** | **p-value** |
| **Neighborhood disadvantage index** | | | | | | | | | |
| The least disadvantaged neighborhoods (<=0) | Ref | Ref | Ref | Ref | Ref | Ref | Ref | Ref | Ref |
| The most disadvantaged neighborhoods (>0) | 1.05 | 0.83, 1.33 | 0.700 | 1.03 | 0.81, 1.30 | 0.800 | 1.11 | 0.84, 1.48 | 0.5 |
| **Age** | 1.05 | 1.04, 1.07 | **<0.001** | 1.05 | 1.04, 1.07 | **<0.001** | 1.05 | 1.04, 1.07 | **<0.001** |
| **Sex** | | | | | | | | | |
| Female | Ref | Ref | Ref | Ref | Ref | Ref | Ref | Ref | Ref |
| Male | 1.19 | 0.93, 1.52 | 0.200 | 1.19 | 0.93, 1.53 | 0.200 | 1.19 | 0.93, 1.53 | 0.2 |
| **Education** | | | | | | | | | |
| Above High School/GED | Ref | Ref | Ref | Ref | Ref | Ref | Ref | Ref | Ref |
| High School/GED | 1.54 | 1.13, 2.10 | **0.006** | 1.54 | 1.12, 2.10 | **0.007** | 1.53 | 1.12, 2.10 | **0.007** |
| Less than High School/GED | 3.12 | 2.18, 4.48 | **<0.001** | 3.02 | 2.10, 4.33 | **<0.001** | 3.03 | 2.11, 4.35 | **<0.001** |
| **Poverty Status** | | | | | | | | | |
| Above Poverty threshold | Ref | Ref | Ref | Ref | Ref | Ref | Ref | Ref | Ref |
| Below Poverty threshold | 1.79 | 1.34, 2.39 | **<0.001** | 1.79 | 1.34, 2.40 | **<0.001** | 1.8 | 1.35, 2.40 | **<0.001** |
| **APOE E4 status** | | | | | | | | | |
| No copies of e4 | Ref | Ref | Ref | Ref | Ref | Ref | Ref | Ref | Ref |
| Any copies of e4 | 1 | 0.79, 1.27 | >0.9 | 1.00 | 0.79, 1.27 | >0.9 | 1 | 0.79, 1.27 | >0.9 |
| **Social Ladder** | 0.97 | 0.91, 1.04 | 0.400 | 0.96 | 0.90, 1.03 | 0.200 | 0.96 | 0.90, 1.03 | **0.3** |
| **Baseline wave** | | | | | | | | | |
| Wave 1 (2008) | Ref | Ref | Ref | Ref | Ref | Ref | Ref | Ref | Ref |
| Wave 2 (2010) | 1.19 | 0.93, 1.52 | 0.200 | 1.18 | 0.92, 1.50 | 0.200 | 1.17 | 0.91, 1.49 | 0.2 |
| **PGS-AD** | | | | | | | | | |
| Below 75% | - | - | - | Ref | Ref | Ref | Ref | Ref | Ref |
| Above 75% | - | - | - | 1.07 | 0.80, 1.43 | 0.700 | 1.28 | 0.83, 1.98 | 0.3 |
| **Neighborhood* PGS-AD** | - | - | - | - | - | - | 0.75 | 0.44, 1.27 | 0.3 |
| RERI: The most disadvantaged neighborhoods*PGS-AD Above 75% | - | - | - | - | - | - | -0.33 | -0.97, 0.31 | |

| | **Dementia vs. Non-dementia, African Ancestry (n=971)** | | | | | | | | |
|---|---|---|---|---|---|---|---|---|---|
| | **Model 1** | | | **Model 2** | | | **Model 3** | | |
| | **HR** | **95% CI** | **p-value** | **HR** | **95% CI** | **p-value** | **HR** | **95% CI** | **p-value** |
| **Neighborhood disadvantage index** | | | | | | | | | |
| The least disadvantaged neighborhoods (<=0) | Ref | Ref | Ref | Ref | Ref | Ref | Ref | Ref | Ref |
| The most disadvantaged neighborhoods (>0) | 1.17 | 0.85, 1.61 | 0.300 | 1.17 | 0.85, 1.61 | 0.300 | 1.21 | 0.84, 1.74 | 0.3 |
| **Age** | 1.09 | 1.07, 1.11 | **<0.001** | 1.09 | 1.07, 1.11 | **<0.001** | 1.09 | 1.07, 1.11 | **<0.001** |
| **Sex** | | | | | | | | | |
| Female | Ref | Ref | Ref | Ref | Ref | Ref | Ref | Ref | Ref |
| Male | 1.32 | 0.96, 1.81 | 0.089 | 1.36 | 0.99, 1.88 | 0.059 | 1.36 | 0.99, 1.87 | 0.06 |
| **Education** | | | | | | | | | |
| Above High School/GED | Ref | Ref | Ref | Ref | Ref | Ref | Ref | Ref | Ref |
| High School/GED | 2.14 | 1.16, 3.95 | **0.016** | 2.19 | 1.18, 4.05 | **0.013** | 2.18 | 1.18, 4.03 | **0.013** |
| Less than High School/GED | 5.51 | 2.96, 10.3 | **<0.001** | 5.63 | 3.01, 10.5 | **<0.001** | 5.63 | 3.01, 10.5 | **<0.001** |
| **Poverty Status** | | | | | | | | | |
| Above Poverty threshold | Ref | Ref | Ref | Ref | Ref | Ref | Ref | Ref | Ref |
| Below Poverty threshold | 1.87 | 1.31, 2.66 | **<0.001** | 1.84 | 1.29, 2.62 | **<0.001** | 1.83 | 1.28, 2.62 | **<0.001** |
| **APOE E4 status** | | | | | | | | | |
| No copies of e4 | Ref | Ref | Ref | Ref | Ref | Ref | Ref | Ref | Ref |
| Any copies of e4 | 1.44 | 1.05, 1.96 | **0.024** | 1.45 | 1.06, 1.99 | **0.020** | 1.45 | 1.06, 1.99 | **0.021** |
| **Social Ladder** | 1.07 | 0.98, 1.16 | 0.150 | 1.06 | 0.98, 1.16 | 0.200 | 1.06 | 0.98, 1.16 | **0.2** |
| **Baseline wave** | | | | | | | | | |
| Wave 1 (2008) | Ref | Ref | Ref | Ref | Ref | Ref | Ref | Ref | Ref |
| Wave 2 (2010) | 1.12 | 0.82, 1.52 | 0.500 | 1.11 | 0.81, 1.52 | 0.500 | 1.11 | 0.81, 1.52 | 0.5 |
| **PGS-AD** | | | | | | | | | |
| Below 75% | - | - | - | Ref | Ref | Ref | Ref | Ref | Ref |
| Above 75% | - | - | - | 0.98 | 0.65, 1.47 | >0.9 | 1.05 | 0.58, 1.90 | 0.9 |
| **Neighborhood* PGS-AD** | - | - | - | - | - | - | 0.88 | 0.42, 1.86 | 0.7 |
| RERI: The most disadvantaged neighborhoods*PGS-AD Above 75% | - | - | - | - | - | - | -0.14 | -0.96, 0.68 | |

**Supplemental Table 4.** Hazard Ratios from survival analysis with additional sensitivity covariates stratified by Ancestry, estimates present the association for each standard deviation increase in the neighborhood disadvantage index with incident cognitive impairment (CIND and dementia), CIND and dementia, relative to normal cognition and non-dementia respectively in the US Health and Retirement Study (2008-2010 Waves).

| | **Cognitive Impairment vs. Normal Cognition, European Ancestry (n=6,035)** | | | | | | | | | **CIND vs. Normal Cognition, European Ancestry (n=5,966)** | | | | | | | | | **Dementia vs. Non-dementia, European Ancestry (n=6,689)** | | | | | | | | |
|---|---|---|---|---|---|---|---|---|---|---|---|---|---|---|---|---|---|---|---|---|---|---|---|---|---|---|---|
| | **Model 1** | | | **Model 2** | | | **Model 3** | | | **Model 1** | | | **Model 2** | | | **Model 3** | | | **Model 1** | | | **Model 2** | | | **Model 3** | | |
| | **HR** | **95% CI** | **p-value** | **HR** | **95% CI** | **p-value** | **HR** | **95% CI** | **p-value** | **HR** | **95% CI** | **p-value** | **HR** | **95% CI** | **p-value** | **HR** | **95% CI** | **p-value** | **HR** | **95% CI** | **p-value** | **HR** | **95% CI** | **p-value** | **HR** | **95% CI** | **p-value** |
| **Neighborhood** | 1.06 | 1.00, 1.12 | **0.040** | 1.06 | 1.00, 1.12 | **0.040** | 1.06 | 1.00, 1.12 | **0.041** | 1.05 | 1.00, 1.11 | 0.058 | 1.05 | 1.00, 1.11 | 0.061 | 1.05 | 1.00, 1.11 | 0.062 | 1.11 | 1.01, 1.22 | **0.038** | 1.10 | 1.00, 1.22 | **0.046** | 1.10 | 1.00, 1.22 | **0.048** |
| **Age** | 1.08 | 1.07, 1.09 | **<0.001** | 1.08 | 1.07, 1.09 | **<0.001** | 1.08 | 1.07, 1.09 | **<0.001** | 1.08 | 1.07, 1.08 | **<0.001** | 1.08 | 1.07, 1.09 | **<0.001** | 1.08 | 1.07, 1.09 | **<0.001** | 1.11 | 1.10, 1.13 | **<0.001** | 1.11 | 1.10, 1.13 | **<0.001** | 1.11 | 1.10, 1.13 | **<0.001** |
| **Sex** | | | | | | | | | | | | | | | | | | | | | | | | | | | |
| Female | Ref | Ref | Ref | Ref | Ref | Ref | Ref | Ref | Ref | Ref | Ref | Ref | Ref | Ref | Ref | Ref | Ref | Ref | Ref | Ref | Ref | Ref | Ref | Ref | Ref | Ref | Ref |
| Male | 1.29 | 1.17, 1.43 | **<0.001** | 1.29 | 1.17, 1.43 | **<0.001** | 1.29 | 1.17, 1.43 | **<0.001** | 1.30 | 1.17, 1.44 | **<0.001** | 1.30 | 1.17, 1.44 | **<0.001** | 1.30 | 1.17, 1.44 | **<0.001** | 1.15 | 0.94, 1.41 | 0.200 | 1.16 | 0.94, 1.42 | 0.200 | 1.16 | 0.94, 1.42 | 0.200 |
| **Education** | | | | | | | | | | | | | | | | | | | | | | | | | | | |
| Above High School/GED | Ref | Ref | Ref | Ref | Ref | Ref | Ref | Ref | Ref | Ref | Ref | Ref | Ref | Ref | Ref | Ref | Ref | Ref | Ref | Ref | Ref | Ref | Ref | Ref | Ref | Ref | Ref |
| High School/GED | 1.59 | 1.41, 1.79 | **<0.001** | 1.59 | 1.41, 1.79 | **<0.001** | 1.59 | 1.41, 1.79 | **<0.001** | 1.62 | 1.43, 1.82 | **<0.001** | 1.62 | 1.44, 1.83 | **<0.001** | 1.62 | 1.44, 1.83 | **<0.001** | 1.63 | 1.26, 2.11 | **<0.001** | 1.64 | 1.27, 2.13 | **<0.001** | 1.64 | 1.27, 2.13 | **<0.001** |
| Less than High School/GED | 2.84 | 2.40, 3.37 | **<0.001** | 2.84 | 2.40, 3.37 | **<0.001** | 2.84 | 2.40, 3.37 | **<0.001** | 2.90 | 2.44, 3.45 | **<0.001** | 2.91 | 2.44, 3.46 | **<0.001** | 2.91 | 2.44, 3.46 | **<0.001** | 4.01 | 2.95, 5.45 | **<0.001** | 4.06 | 2.99, 5.52 | **<0.001** | 4.06 | 2.98, 5.52 | **<0.001** |
| **Poverty Status (Below)** | | | | | | | | | | | | | | | | | | | | | | | | | | | |
| Above Poverty threshold | Ref | Ref | Ref | Ref | Ref | Ref | Ref | Ref | Ref | Ref | Ref | Ref | Ref | Ref | Ref | Ref | Ref | Ref | Ref | Ref | Ref | Ref | Ref | Ref | Ref | Ref | Ref |
| Below Poverty threshold | 1.16 | 0.89, 1.50 | 0.300 | 1.15 | 0.89, 1.49 | 0.300 | 1.15 | 0.89, 1.49 | 0.300 | 1.17 | 0.90, 1.52 | 0.200 | 1.17 | 0.90, 1.52 | 0.200 | 1.17 | 0.90, 1.52 | 0.200 | 1.85 | 1.24, 2.75 | **0.003** | 1.89 | 1.27, 2.81 | **0.002** | 1.89 | 1.27, 2.82 | **0.002** |
| **APOE E4 status (No copy)** | | | | | | | | | | | | | | | | | | | | | | | | | | | |
| No copies of e4 | Ref | Ref | Ref | Ref | Ref | Ref | Ref | Ref | Ref | Ref | Ref | Ref | Ref | Ref | Ref | Ref | Ref | Ref | Ref | Ref | Ref | Ref | Ref | Ref | Ref | Ref | Ref |
| Any copies of e4 | 1.44 | 1.30, 1.60 | **<0.001** | 1.42 | 1.28, 1.58 | **<0.001** | 1.42 | 1.28, 1.58 | **<0.001** | 1.45 | 1.30, 1.61 | **<0.001** | 1.43 | 1.29, 1.60 | **<0.001** | 1.43 | 1.29, 1.60 | **<0.001** | 2.07 | 1.70, 2.52 | **<0.001** | 2.06 | 1.69, 2.51 | **<0.001** | 2.06 | 1.69, 2.50 | **<0.001** |
| **Social Ladder** | 0.95 | 0.92, 0.98 | **0.001** | 0.95 | 0.92, 0.98 | **0.001** | 0.95 | 0.92, 0.98 | **0.001** | 0.95 | 0.92, 0.98 | **<0.001** | 0.95 | 0.92, 0.98 | **0.001** | 0.95 | 0.92, 0.98 | **<0.001** | 1.00 | 0.94, 1.06 | >0.9 | 1.00 | 0.94, 1.06 | >0.9 | 1.00 | 0.94, 1.06 | >0.9 |
| **Baseline wave** | | | | | | | | | | | | | | | | | | | | | | | | | | | |
| Wave 1 (2008) | Ref | Ref | Ref | Ref | Ref | Ref | Ref | Ref | Ref | Ref | Ref | Ref | Ref | Ref | Ref | Ref | Ref | Ref | Ref | Ref | Ref | Ref | Ref | Ref | Ref | Ref | Ref |
| Wave 2 (2010) | 0.86 | 0.78, 0.94 | **0.001** | 0.86 | 0.78, 0.95 | **0.002** | 0.86 | 0.78, 0.95 | **0.002** | 0.86 | 0.78, 0.95 | **0.002** | 0.86 | 0.78, 0.95 | **0.003** | 0.86 | 0.78, 0.95 | **0.003** | 0.80 | 0.67, 0.97 | **0.024** | 0.80 | 0.67, 0.97 | **0.024** | 0.8 | 0.67, 0.97 | **0.024** |
| **PGS-AD** | - | - | - | 1.11 | 1.05, 1.16 | **<0.001** | 1.10 | 1.05, 1.16 | **<0.001** | - | - | - | 1.10 | 1.05, 1.16 | **<0.001** | 1.10 | 1.05, 1.16 | **<0.001** | - | - | - | 1.05 | 0.95, 1.16 | 0.300 | 1.05 | 0.95, 1.16 | 0.300 |
| **Neighborhood* PGS-AD** | - | - | - | - | - | - | 0.99 | 0.94, 1.05 | 0.800 | - | - | - | - | - | - | 1.00 | 0.94, 1.05 | 0.800 | - | - | - | - | - | - | 0.99 | 0.90, 1.09 | 0.800 |
| **Smoking status** | | | | | | | | | | | | | | | | | | | | | | | | | | | |
| Never Smoker | Ref | Ref | Ref | Ref | Ref | Ref | Ref | Ref | Ref | Ref | Ref | Ref | Ref | Ref | Ref | Ref | Ref | Ref | Ref | Ref | Ref | Ref | Ref | Ref | Ref | Ref | Ref |
| Current Smoker | 1.28 | 1.08, 1.52 | **0.005** | 1.29 | 1.08, 1.53 | **0.004** | 1.29 | 1.08, 1.53 | **0.004** | 1.29 | 1.08, 1.54 | **0.004** | 1.29 | 1.09, 1.54 | **0.004** | 1.29 | 1.09, 1.54 | **0.004** | 1.06 | 0.72, 1.56 | 0.800 | 1.05 | 0.71, 1.54 | 0.800 | 1.05 | 0.71, 1.54 | 0.800 |
| Former Smoke | 1.07 | 0.97, 1.19 | 0.200 | 1.06 | 0.96, 1.18 | 0.300 | 1.06 | 0.96, 1.18 | 0.300 | 1.07 | 0.96, 1.19 | 0.200 | 1.06 | 0.95, 1.17 | 0.300 | 1.06 | 0.95, 1.17 | 0.300 | 1.01 | 0.82, 1.24 | >0.9 | 1.00 | 0.82, 1.23 | >0.9 | 1.00 | 0.82, 1.23 | >0.9 |
| **Drinking (# drinks/day when drinks)** | 1.01 | 0.97, 1.05 | 0.800 | 1.01 | 0.97, 1.05 | 0.700 | 1.01 | 0.97, 1.05 | 0.700 | 1.01 | 0.97, 1.05 | 0.700 | 1.01 | 0.97, 1.05 | 0.600 | 1.01 | 0.97, 1.05 | 0.600 | 1.01 | 0.92, 1.11 | 0.800 | 1.01 | 0.92, 1.11 | 0.800 | 1.01 | 0.92, 1.11 | 0.800 |
| **Depression** | 1.09 | 1.06, 1.12 | **<0.001** | 1.09 | 1.06, 1.12 | **<0.001** | 1.09 | 1.06, 1.12 | **<0.001** | 1.09 | 1.06, 1.12 | **<0.001** | 1.09 | 1.06, 1.12 | **<0.001** | 1.09 | 1.06, 1.12 | **<0.001** | 1.11 | 1.05, 1.17 | **<0.001** | 1.11 | 1.06, 1.17 | **<0.001** | 1.11 | 1.06, 1.17 | **<0.001** |
| **BMI** | 0.99 | 0.98, 1.00 | 0.200 | 1.00 | 0.99, 1.00 | 0.300 | 1.00 | 0.99, 1.00 | 0.300 | 1.00 | 0.99, 1.00 | 0.300 | 1.00 | 0.99, 1.01 | 0.400 | 1.00 | 0.99, 1.01 | 0.400 | 0.97 | 0.95, 0.99 | **0.005** | 0.97 | 0.95, 0.99 | **0.006** | 0.97 | 0.95, 0.99 | **0.006** |
| **Ever have Diabetes** | | | | | | | | | | | | | | | | | | | | | | | | | | | |
| No | Ref | Ref | Ref | Ref | Ref | Ref | Ref | Ref | Ref | Ref | Ref | Ref | Ref | Ref | Ref | Ref | Ref | Ref | Ref | Ref | Ref | Ref | Ref | Ref | Ref | Ref | Ref |
| Yes | 1.14 | 1.00, 1.30 | 0.059 | 1.13 | 0.98, 1.29 | 0.086 | 1.13 | 0.98, 1.29 | 0.086 | 1.16 | 1.02, 1.33 | **0.029** | 1.15 | 1.00, 1.32 | **0.046** | 1.15 | 1.00, 1.32 | 0.046 | 1.06 | 0.80, 1.39 | 0.700 | 1.05 | 0.80, 1.38 | 0.700 | 1.05 | 0.80, 1.38 | 0.700 |
| **Chronic Condition** | | | | | | | | | | | | | | | | | | | | | | | | | | | |
| None | Ref | Ref | Ref | Ref | Ref | Ref | Ref | Ref | Ref | Ref | Ref | Ref | Ref | Ref | Ref | Ref | Ref | Ref | Ref | Ref | Ref | Ref | Ref | Ref | Ref | Ref | Ref |
| 1 - 2 | 0.99 | 0.84, 1.17 | 0.900 | 0.99 | 0.83, 1.17 | 0.900 | 0.99 | 0.83, 1.17 | 0.900 | 1.00 | 0.84, 1.19 | >0.9 | 1.00 | 0.84, 1.18 | >0.9 | 1.00 | 0.84, 1.18 | >0.9 | 0.78 | 0.56, 1.07 | 0.130 | 0.78 | 0.56, 1.07 | 0.120 | 0.78 | 0.56, 1.07 | 0.120 |
| >= 3 | 1.16 | 0.96, 1.39 | 0.120 | 1.16 | 0.97, 1.40 | 0.110 | 1.16 | 0.97, 1.40 | 0.110 | 1.16 | 0.96, 1.39 | 0.130 | 1.16 | 0.96, 1.40 | 0.110 | 1.16 | 0.96, 1.40 | 0.110 | 0.82 | 0.58, 1.17 | 0.300 | 0.82 | 0.58, 1.16 | 0.300 | 0.82 | 0.58, 1.16 | 0.300 |
| | **Cognitive Impairment vs. Normal Cognition, African Ancestry (n=696)** | | | | | | | | | **CIND vs. Normal Cognition, African Ancestry (n=688)** | | | | | | | | | **Dementia vs. Non-dementia, African Ancestry (n=960)** | | | | | | | | |
| | **Model 1** | | | **Model 2** | | | **Model 3** | | | **Model 1** | | | **Model 2** | | | **Model 3** | | | **Model 1** | | | **Model 2** | | | **Model 3** | | |
| | **HR** | **95% CI** | **p-value** | **HR** | **95% CI** | **p-value** | **HR** | **95% CI** | **p-value** | **HR** | **95% CI** | **p-value** | **HR** | **95% CI** | **p-value** | **HR** | **95% CI** | **p-value** | **HR** | **95% CI** | **p-value** | **HR** | **95% CI** | **p-value** | **HR** | **95% CI** | **p-value** |
| **Neighborhood** | 1.02 | 0.92, 1.14 | 0.700 | 1.02 | 0.92, 1.14 | 0.700 | 1.03 | 0.92, 1.15 | 0.600 | 1.02 | 0.91, 1.14 | 0.700 | 1.02 | 0.91, 1.14 | 0.800 | 1.02 | 0.91, 1.15 | 0.700 | 1.04 | 0.90, 1.20 | 0.600 | 1.05 | 0.91, 1.21 | 0.500 | 1.05 | 0.91, 1.22 | 0.500 |
| **Age** | 1.05 | 1.04, 1.07 | **<0.001** | 1.06 | 1.04, 1.07 | **<0.001** | 1.06 | 1.04, 1.07 | **<0.001** | 1.06 | 1.04, 1.07 | **<0.001** | 1.06 | 1.04, 1.07 | **<0.001** | 1.06 | 1.04, 1.07 | **<0.001** | 1.09 | 1.07, 1.11 | **<0.001** | 1.09 | 1.07, 1.12 | **<0.001** | 1.09 | 1.07, 1.11 | **<0.001** |
| **Sex** | | | | | | | | | | | | | | | | | | | | | | | | | | | |
| Female | Ref | Ref | Ref | Ref | Ref | Ref | Ref | Ref | Ref | Ref | Ref | Ref | Ref | Ref | Ref | Ref | Ref | Ref | Ref | Ref | Ref | Ref | Ref | Ref | Ref | Ref | Ref |
| Male | 1.18 | 0.91, 1.52 | 0.200 | 1.18 | 0.92, 1.53 | 0.200 | 1.19 | 0.92, 1.54 | 0.200 | 1.19 | 0.92, 1.53 | 0.200 | 1.18 | 0.91, 1.54 | 0.200 | 1.19 | 0.92, 1.54 | 0.200 | 1.25 | 0.89, 1.76 | 0.200 | 1.29 | 0.91, 1.82 | 0.150 | 1.29 | 0.91, 1.82 | 0.150 |
| **Education** | | | | | | | | | | | | | | | | | | | | | | | | | | | |
| Above High School/GED | Ref | Ref | Ref | Ref | Ref | Ref | Ref | Ref | Ref | Ref | Ref | Ref | Ref | Ref | Ref | Ref | Ref | Ref | Ref | Ref | Ref | Ref | Ref | Ref | Ref | Ref | Ref |
| High School/GED | 1.49 | 1.10, 2.02 | **0.011** | 1.47 | 1.08, 2.00 | **0.014** | 1.47 | 1.08, 2.00 | **0.014** | 1.51 | 1.11, 2.07 | **0.010** | 1.49 | 1.09, 2.05 | **0.013** | 1.49 | 1.09, 2.05 | **0.013** | 2.08 | 1.12, 3.86 | **0.020** | 2.19 | 1.18, 4.06 | **0.013** | 2.19 | 1.18, 4.07 | **0.013** |
| Less than High School/GED | 3.00 | 2.10, 4.29 | **<0.001** | 2.87 | 2.01, 4.11 | **<0.001** | 2.90 | 2.02, 4.15 | **<0.001** | 3.11 | 2.16, 4.47 | **<0.001** | 2.95 | 2.05, 4.24 | **<0.001** | 2.96 | 2.06, 4.27 | **<0.001** | 5.18 | 2.77, 9.68 | **<0.001** | 5.41 | 2.88, 10.2 | **<0.001** | 5.41 | 2.88, 10.2 | **<0.001** |
| **Poverty Status** | | | | | | | | | | | | | | | | | | | | | | | | | | | |
| Above Poverty threshold | Ref | Ref | Ref | Ref | Ref | Ref | Ref | Ref | Ref | Ref | Ref | Ref | Ref | Ref | Ref | Ref | Ref | Ref | Ref | Ref | Ref | Ref | Ref | Ref | Ref | Ref | Ref |
| Below Poverty threshold | 1.51 | 1.12, 2.03 | **0.007** | 1.52 | 1.13, 2.06 | **0.006** | 1.54 | 1.14, 2.08 | **0.005** | 1.50 | 1.11, 2.02 | **0.008** | 1.50 | 1.11, 2.03 | **0.008** | 1.52 | 1.12, 2.06 | **0.006** | 1.56 | 1.07, 2.27 | **0.021** | 1.52 | 1.04, 2.22 | **0.029** | 1.53 | 1.04, 2.23 | **0.029** |
| **APOE E4 status** | | | | | | | | | | | | | | | | | | | | | | | | | | | |
| No copies of e4 | Ref | Ref | Ref | Ref | Ref | Ref | Ref | Ref | Ref | Ref | Ref | Ref | Ref | Ref | Ref | Ref | Ref | Ref | Ref | Ref | Ref | Ref | Ref | Ref | Ref | Ref | Ref |
| Any copies of e4 | 0.94 | 0.74, 1.19 | 0.600 | 0.94 | 0.74, 1.19 | 0.600 | 0.92 | 0.72, 1.18 | 0.5 | 0.92 | 0.72, 1.17 | 0.500 | 0.92 | 0.72, 1.18 | 0.500 | 0.91 | 0.71, 1.16 | 0.400 | 1.36 | 0.99, 1.88 | 0.058 | 1.38 | 1.00, 1.91 | **0.048** | 1.35 | 0.98, 1.87 | **0.067** |
| **Social Ladder** | 0.98 | 0.92, 1.05 | 0.600 | 0.97 | 0.91, 1.04 | 0.400 | 0.97 | 0.91, 1.04 | 0.400 | 0.99 | 0.92, 1.06 | 0.700 | 0.98 | 0.92, 1.05 | 0.500 | 0.98 | 0.92, 1.05 | 0.600 | 1.08 | 0.99, 1.18 | 0.083 | 1.08 | 0.99, 1.17 | 0.100 | 1.08 | 0.99, 1.18 | 0.086 |
| **Baseline wave** | | | | | | | | | | | | | | | | | | | | | | | | | | | |
| Wave 1 (2008) | Ref | Ref | Ref | Ref | Ref | Ref | Ref | Ref | Ref | Ref | Ref | Ref | Ref | Ref | Ref | Ref | Ref | Ref | Ref | Ref | Ref | Ref | Ref | Ref | Ref | Ref | Ref |
| Wave 2 (2010) | 1.17 | 0.92, 1.49 | 0.200 | 1.140 | 0.90, 1.46 | 0.300 | 1.14 | 0.90, 1.45 | 0.3 | 1.18 | 0.92, 1.50 | 0.200 | 1.150 | 0.90, 1.47 | 0.300 | 1.15 | 0.90, 1.47 | 0.3 | 1.06 | 0.78, 1.46 | 0.700 | 1.030 | 0.75, 1.42 | 0.900 | 1.03 | 0.75, 1.43 | 0.8 |
| **PGS-AD** | - | - | - | 1.130 | 0.94, 1.34 | 0.200 | 1.15 | 0.96, 1.37 | 0.140 | - | - | - | 1.110 | 0.93, 1.33 | 0.200 | 1.13 | 0.94, 1.35 | 0.200 | - | - | - | 1.110 | 0.87, 1.42 | 0.400 | 1.14 | 0.89, 1.47 | 0.3 |
| **Neighborhood* PGS-AD** | - | - | - | - | - | - | 0.94 | 0.82, 1.07 | 0.400 | - | - | - | - | - | - | 0.94 | 0.83, 1.08 | 0.400 | - | - | - | - | - | - | 0.91 | 0.77, 1.08 | 0.3 |
| **Smoking status** | | | | | | | | | | | | | | | | | | | | | | | | | | | |
| Never Smoker | Ref | Ref | Ref | Ref | Ref | Ref | Ref | Ref | Ref | Ref | Ref | Ref | Ref | Ref | Ref | Ref | Ref | Ref | Ref | Ref | Ref | Ref | Ref | Ref | Ref | Ref | Ref |

| | | | | | | | | | | | | | | | | | | | | | | | | | | | |
|---|---|---|---|---|---|---|---|---|---|---|---|---|---|---|---|---|---|---|---|---|---|---|---|---|---|---|---|
| Current Smoker | 1.23 | 0.88, 1.72 | 0.200 | 1.24 | 0.89, 1.74 | 0.200 | 1.24 | 0.89, 1.74 | 0.2 | 1.25 | 0.89, 1.75 | 0.200 | 1.27 | 0.90, 1.78 | 0.200 | 1.27 | 0.90, 1.78 | 0.200 | 1.40 | 0.84, 2.35 | 0.200 | 1.40 | 0.83, 2.36 | 0.200 | 1.41 | 0.84, 2.39 | 0.2 |
| Former Smoke | 0.95 | 0.73, 1.24 | 0.700 | 0.99 | 0.76, 1.29 | >0.9 | 0.99 | 0.76, 1.29 | >0.9 | 0.92 | 0.71, 1.20 | 0.500 | 0.95 | 0.73, 1.24 | 0.700 | 0.95 | 0.72, 1.24 | 0.700 | 1.42 | 1.00, 2.02 | **0.050** | 1.42 | 1.00, 2.03 | 0.053 | 1.42 | 0.99, 2.02 | 0.056 |
| **Drinking (# drinks/day when drinks)** | 1.03 | 0.94, 1.12 | 0.600 | 1.03 | 0.94, 1.12 | 0.600 | 1.02 | 0.94, 1.12 | 0.6 | 1.03 | 0.94, 1.13 | 0.500 | 1.03 | 0.94, 1.13 | 0.500 | 1.03 | 0.94, 1.13 | 0.500 | 0.99 | 0.87, 1.14 | >0.9 | 0.99 | 0.86, 1.13 | 0.900 | 0.99 | 0.86, 1.13 | 0.9 |
| **Depression** | 1.07 | 1.01, 1.14 | **0.021** | 1.08 | 1.02, 1.14 | **0.014** | 1.08 | 1.02, 1.14 | **0.014** | 1.08 | 1.01, 1.14 | **0.018** | 1.08 | 1.02, 1.15 | **0.012** | 1.08 | 1.02, 1.15 | **0.012** | 1.14 | 1.06, 1.23 | **<0.001** | 1.15 | 1.07, 1.23 | **<0.001** | 1.15 | 1.07, 1.24 | **<0.001** |
| **BMI** | 0.99 | 0.97, 1.01 | 0.400 | 0.99 | 0.97, 1.01 | 0.400 | 0.99 | 0.97, 1.01 | 0.4 | 0.99 | 0.97, 1.01 | 0.500 | 0.99 | 0.97, 1.01 | 0.500 | 0.99 | 0.97, 1.01 | 0.500 | 0.99 | 0.97, 1.02 | 0.500 | 0.99 | 0.97, 1.02 | 0.400 | 0.99 | 0.96, 1.01 | 0.4 |
| **Ever have Diabetes** | | | | | | | | | | | | | | | | | | | | | | | | | | | |
| No | Ref | Ref | Ref | Ref | Ref | Ref | Ref | Ref | Ref | Ref | Ref | Ref | Ref | Ref | Ref | Ref | Ref | Ref | Ref | Ref | Ref | Ref | Ref | Ref | Ref | Ref | Ref |
| Yes | 1.13 | 0.84, 1.52 | 0.400 | 1.16 | 0.86, 1.57 | 0.300 | 1.15 | 0.85, 1.56 | 0.400 | 1.14 | 0.84, 1.54 | 0.400 | 1.16 | 0.86, 1.58 | 0.300 | 1.16 | 0.85, 1.57 | 0.300 | 0.96 | 0.65, 1.43 | 0.900 | 1.01 | 0.67, 1.50 | >0.9 | 1.01 | 0.67, 1.50 | >0.9 |
| **Chronic Condition** | | | | | | | | | | | | | | | | | | | | | | | | | | | |
| None | Ref | Ref | Ref | Ref | Ref | Ref | Ref | Ref | Ref | Ref | Ref | Ref | Ref | Ref | Ref | Ref | Ref | Ref | Ref | Ref | Ref | Ref | Ref | Ref | Ref | Ref | Ref |
| 1 - 2 | 0.70 | 0.49, 1.01 | 0.059 | 0.71 | 0.49, 1.03 | 0.071 | 0.71 | 0.49, 1.03 | 0.075 | 0.68 | 0.47, 0.99 | **0.043** | 0.69 | 0.47, 1.00 | **0.050** | 0.69 | 0.47, 1.01 | 0.053 | 0.84 | 0.49, 1.44 | 0.500 | 0.84 | 0.49, 1.44 | 0.500 | 0.82 | 0.48, 1.42 | 0.5 |
| >= 3 | 0.96 | 0.63, 1.47 | 0.900 | 0.94 | 0.61, 1.45 | 0.800 | 0.95 | 0.62, 1.47 | 0.8 | 0.96 | 0.62, 1.47 | 0.800 | 0.94 | 0.61, 1.46 | 0.800 | 0.96 | 0.62, 1.48 | 0.800 | 0.82 | 0.45, 1.49 | 0.500 | 0.78 | 0.43, 1.44 | 0.400 | 0.78 | 0.43, 1.44 | 0.4 |

Each model further adjusted for Smoking, Alcohol Consumption, BMI, Diabetes, Chronic Conditions, and Depression.

**Supplemental Table 5.** Hazard Ratios from survival analysis using individual neighborhood factor as exposure, stratified by Ancestry: risk of incident cognitive impairment (CIND and dementia), CIND and dementia, relative to normal cognition and non-dementia in the US Health and Retirement Study (2008-2010 Waves).

| | Cognitive Impairment vs. Normal Cognition, European Ancestry (n=6,123) | | | | | | | | | CIND vs. Normal Cognition, European Ancestry (n=6,051) | | | | | | | | | Dementia vs. Non-dementia, European Ancestry (n=6,789) | | | | | | | | |
|---|---|---|---|---|---|---|---|---|---|---|---|---|---|---|---|---|---|---|---|---|---|---|---|---|---|---|---|
| | Model 1 | | | Model 2 | | | Model 3 | | | Model 1 | | | Model 2 | | | Model 3 | | | Model 1 | | | Model 2 | | | Model 3 | | |
| | HR | 95% CI | p-value | HR | 95% CI | p-value | HR | 95% CI | p-value | HR | 95% CI | p-value | HR | 95% CI | p-value | HR | 95% CI | p-value | HR | 95% CI | p-value | HR | 95% CI | p-value | HR | 95% CI | p-value |
| **Neighborhood Safety** | 1.12 | 1.07, 1.18 | **<0.001** | 1.12 | 1.06, 1.18 | **<0.001** | 1.12 | 1.06, 1.18 | **<0.001** | 1.12 | 1.06, 1.18 | **<0.001** | 1.12 | 1.06, 1.18 | **<0.001** | 1.12 | 1.06, 1.18 | **<0.001** | 1.16 | 1.06, 1.28 | **0.002** | 1.16 | 1.05, 1.27 | **0.003** | 1.16 | 1.05, 1.27 | **0.003** |
| PGS-AD | - | - | - | 1.10 | 1.05, 1.16 | **<0.001** | 1.10 | 1.05, 1.15 | **<0.001** | - | - | - | 1.10 | 1.05, 1.16 | **<0.001** | 1.10 | 1.04, 1.15 | **<0.001** | - | - | - | 1.05 | 0.95, 1.15 | 0.300 | 1.05 | 0.95, 1.15 | 0.3 |
| Neighborhood Safety*PGS-AD | - | - | - | - | - | - | 0.96 | 0.91, 1.01 | 0.13 | - | - | - | - | - | - | 0.96 | 0.91, 1.01 | 0.15 | - | - | - | - | - | - | 0.97 | 0.88, 1.07 | 0.5 |
| **Neighborhood Trust** | 1.06 | 1.01, 1.12 | **0.029** | 1.07 | 1.01, 1.13 | **0.024** | 1.07 | 1.01, 1.13 | **0.024** | 1.06 | 1.00, 1.12 | **0.041** | 1.06 | 1.00, 1.12 | **0.036** | 1.06 | 1.00, 1.12 | **0.037** | 1.09 | 0.98, 1.20 | 0.100 | 1.08 | 0.98, 1.20 | 0.120 | 1.08 | 0.98, 1.20 | 0.130 |
| PGS-AD | - | - | - | 1.10 | 1.05, 1.16 | **<0.001** | 1.10 | 1.05, 1.16 | **<0.001** | - | - | - | 1.10 | 1.05, 1.16 | **<0.001** | 1.10 | 1.05, 1.16 | **<0.001** | - | - | - | 1.05 | 0.96, 1.16 | 0.300 | 1.05 | 0.95, 1.16 | 0.4 |
| Neighborhood Trust * PGS-AD | - | - | - | - | - | - | 0.99 | 0.94, 1.05 | 0.7 | - | - | - | - | - | - | 0.99 | 0.94, 1.05 | 0.8 | - | - | - | - | - | - | 0.98 | 0.89, 1.08 | 0.7 |
| **Neighborhood Friendly** | 1.07 | 1.01, 1.12 | **0.017** | 1.07 | 1.01, 1.12 | **0.015** | 1.07 | 1.01, 1.12 | **0.017** | 1.06 | 1.01, 1.12 | **0.031** | 1.06 | 1.01, 1.12 | **0.029** | 1.06 | 1.01, 1.12 | **0.032** | 1.09 | 0.99, 1.19 | 0.089 | 1.08 | 0.98, 1.19 | 0.110 | 1.08 | 0.98, 1.19 | 0.12 |
| PGS-AD | - | - | - | 1.10 | 1.05, 1.16 | **<0.001** | 1.10 | 1.05, 1.15 | **<0.001** | - | - | - | 1.10 | 1.05, 1.16 | **<0.001** | 1.10 | 1.05, 1.15 | **<0.001** | - | - | - | 1.05 | 0.96, 1.16 | 0.300 | 1.05 | 0.95, 1.15 | 0.3 |
| Neighborhood Friendly * PGS-AD | - | - | - | - | - | - | 0.97 | 0.93, 1.02 | 0.3 | - | - | - | - | - | - | 0.98 | 0.93, 1.03 | 0.4 | - | - | - | - | - | - | 0.97 | 0.88, 1.06 | 0.5 |
| **Neighborhood Vandalism** | 1.06 | 1.00, 1.11 | **0.040** | 1.06 | 1.00, 1.12 | **0.033** | 1.06 | 1.00, 1.12 | **0.033** | 1.05 | 1.00, 1.11 | 0.066 | 1.05 | 1.00, 1.11 | 0.057 | 1.05 | 1.00, 1.11 | 0.058 | 1.09 | 0.99, 1.21 | 0.068 | 1.09 | 0.99, 1.20 | 0.079 | 1.09 | 0.99, 1.20 | 0.081 |
| PGS-AD | - | - | - | 1.10 | 1.05, 1.16 | **<0.001** | 1.11 | 1.05, 1.16 | **<0.001** | - | - | - | 1.10 | 1.05, 1.16 | **<0.001** | 1.1 | 1.05, 1.16 | **<0.001** | - | - | - | 1.05 | 0.96, 1.16 | 0.300 | 1.05 | 0.95, 1.16 | 0.3 |
| Neighborhood Vandalism* PGS-AD | - | - | - | - | - | - | 1.02 | 0.97, 1.07 | 0.500 | - | - | - | - | - | - | 1.02 | 0.96, 1.07 | 0.6 | - | - | - | - | - | - | 0.98 | 0.90, 1.08 | 0.7 |
| **Neighborhood Belonging** | 1.06 | 1.01, 1.11 | **0.030** | 1.06 | 1.00, 1.11 | **0.034** | 1.06 | 1.00, 1.11 | **0.038** | 1.05 | 1.00, 1.11 | **0.049** | 1.05 | 1.00, 1.11 | 0.058 | 1.05 | 1.00, 1.11 | 0.066 | 1.1 | 1.00, 1.21 | **0.044** | 1.1 | 1.00, 1.21 | **0.050** | 1.10 | 1.00, 1.21 | 0.052 |
| PGS-AD | - | - | - | 1.10 | 1.05, 1.16 | **<0.001** | 1.10 | 1.05, 1.15 | **<0.001** | - | - | - | 1.10 | 1.05, 1.16 | **<0.001** | 1.10 | 1.04, 1.15 | **<0.001** | - | - | - | 1.05 | 0.95, 1.16 | 0.300 | 1.05 | 0.95, 1.16 | 0.300 |
| Neighborhood Belonging * PGS-AD | - | - | - | - | - | - | 0.96 | 0.91, 1.01 | 0.11 | - | - | - | - | - | - | 0.95 | 0.91, 1.01 | 0.082 | - | - | - | - | - | - | 0.99 | 0.89, 1.09 | 0.800 |
| **Neighborhood Cleanness** | 1.06 | 1.01, 1.12 | **0.024** | 1.06 | 1.01, 1.12 | **0.032** | 1.06 | 1.01, 1.12 | **0.032** | 1.06 | 1.00, 1.12 | **0.041** | 1.06 | 1.00, 1.11 | 0.055 | 1.05 | 1.00, 1.11 | 0.056 | 1.11 | 1.01, 1.22 | **0.038** | 1.10 | 1.00, 1.21 | 0.055 | 1.10 | 1.00, 1.21 | 0.057 |
| PGS-AD | - | - | - | 1.10 | 1.05, 1.16 | **<0.001** | 1.10 | 1.05, 1.16 | **<0.001** | - | - | - | 1.10 | 1.05, 1.16 | **<0.001** | 1.1 | 1.05, 1.16 | **<0.001** | - | - | - | 1.05 | 0.95, 1.16 | 0.300 | 1.05 | 0.95, 1.15 | 0.3 |
| Neighborhood Cleanness * PGS-AD | - | - | - | - | - | - | 1 | 0.95, 1.05 | >0.9 | - | - | - | - | - | - | 1 | 0.95, 1.06 | 0.9 | - | - | - | - | - | - | 0.98 | 0.89, 1.08 | 0.7 |
| **Neighborhood Vacant** | 1.04 | 0.99, 1.09 | 0.140 | 1.04 | 0.99, 1.09 | 0.200 | 1.04 | 0.98, 1.09 | 0.2 | 1.04 | 0.98, 1.09 | 0.200 | 1.04 | 0.98, 1.09 | 0.200 | 1.03 | 0.98, 1.09 | 0.2 | 1.1 | 1.00, 1.21 | 0.052 | 1.1 | 1.00, 1.21 | 0.059 | 1.10 | 1.00, 1.21 | 0.059 |
| PGS-AD | - | - | - | 1.10 | 1.05, 1.16 | **<0.001** | 1.11 | 1.06, 1.17 | **<0.001** | - | - | - | 1.10 | 1.05, 1.16 | **<0.001** | 1.11 | 1.06, 1.17 | **<0.001** | - | - | - | 1.05 | 0.95, 1.16 | 0.300 | 1.05 | 0.95, 1.16 | 0.3 |
| Neighborhood Vacant * PGS-AD | - | - | - | - | - | - | 1.06 | 1.01, 1.12 | **0.023** | - | - | - | - | - | - | 1.06 | 1.01, 1.12 | **0.022** | - | - | - | - | - | - | 1.01 | 0.92, 1.12 | 0.8 |

| | Cognitive Impairment vs. Normal Cognition, African Ancestry (n=703) | | | | | | | | | CIND vs. Normal Cognition, African Ancestry (n=695) | | | | | | | | | Dementia vs. Non-dementia, African Ancestry (n=971) | | | | | | | | |
|---|---|---|---|---|---|---|---|---|---|---|---|---|---|---|---|---|---|---|---|---|---|---|---|---|---|---|---|
| | Model 1 | | | Model 2 | | | Model 3 | | | Model 1 | | | Model 2 | | | Model 3 | | | Model 1 | | | Model 2 | | | Model 3 | | |
| | HR | 95% CI | p-value | HR | 95% CI | p-value | HR | 95% CI | p-value | HR | 95% CI | p-value | HR | 95% CI | p-value | HR | 95% CI | p-value | HR | 95% CI | p-value | HR | 95% CI | p-value | HR | 95% CI | p-value |
| **Neighborhood Safety** | 1.03 | 0.92, 1.14 | 0.600 | 1.03 | 0.93, 1.15 | 0.600 | 1.04 | 0.93, 1.16 | 0.5 | 1.03 | 0.92, 1.14 | 0.600 | 1.04 | 0.93, 1.15 | 0.500 | 1.04 | 0.93, 1.16 | 0.500 | 1.07 | 0.93, 1.22 | 0.400 | 1.07 | 0.93, 1.24 | 0.300 | 1.08 | 0.94, 1.25 | 0.300 |
| PGS-AD | - | - | - | 1.13 | 0.95, 1.35 | 0.200 | 1.15 | 0.96, 1.37 | 0.13 | - | - | - | 1.11 | 0.93, 1.33 | 0.200 | 1.12 | 0.94, 1.34 | 0.2 | - | - | - | 1.10 | 0.86, 1.39 | 0.500 | 1.14 | 0.89, 1.46 | 0.3 |
| Neighborhood Safety*PGS-AD | - | - | - | - | - | - | 0.95 | 0.84, 1.07 | 0.4 | - | - | - | - | - | - | 0.96 | 0.85, 1.08 | 0.5 | - | - | - | - | - | - | 0.87 | 0.74, 1.03 | 0.11 |
| **Neighborhood Trust** | 1.01 | 0.91, 1.13 | 0.900 | 0.99 | 0.89, 1.11 | 0.900 | 1.01 | 0.91, 1.13 | 0.800 | 0.99 | 0.89, 1.11 | >0.9 | 0.97 | 0.87, 1.09 | 0.600 | 1.00 | 0.89, 1.12 | >0.9 | 1.06 | 0.93, 1.22 | 0.400 | 1.06 | 0.92, 1.22 | 0.400 | 1.07 | 0.93, 1.23 | 0.300 |
| PGS-AD | - | - | - | 1.14 | 0.95, 1.35 | 0.200 | 1.18 | 0.99, 1.42 | 0.065 | - | - | - | 1.12 | 0.94, 1.33 | 0.200 | 1.18 | 0.98, 1.41 | 0.073 | - | - | - | 1.10 | 0.86, 1.39 | 0.400 | 1.12 | 0.87, 1.44 | 0.4 |
| Neighborhood Trust * PGS-AD | - | - | - | - | - | - | 0.88 | 0.77, 1.01 | 0.064 | - | - | - | - | - | - | 0.86 | 0.75, 0.98 | **0.025** | - | - | - | - | - | - | 0.95 | 0.81, 1.13 | 0.6 |
| **Neighborhood Friendly** | 1.11 | 1.00, 1.23 | 0.051 | 1.12 | 1.00, 1.24 | **0.041** | 1.12 | 1.00, 1.24 | **0.046** | 1.11 | 1.00, 1.24 | **0.050** | 1.12 | 1.01, 1.25 | **0.039** | 1.12 | 1.00, 1.25 | **0.048** | 1.01 | 0.88, 1.17 | 0.800 | 1.02 | 0.89, 1.17 | 0.800 | 1.08 | 0.94, 1.25 | 0.3 |
| PGS-AD | - | - | - | 1.13 | 0.95, 1.35 | 0.200 | 1.13 | 0.94, 1.35 | 0.2 | - | - | - | 1.11 | 0.93, 1.33 | 0.200 | 1.11 | 0.93, 1.33 | 0.3 | - | - | - | 1.09 | 0.86, 1.39 | 0.500 | 1.14 | 0.89, 1.46 | 0.3 |
| Neighborhood Friendly * PGS-AD | - | - | - | - | - | - | 1.00 | 0.87, 1.15 | >0.9 | - | - | - | - | - | - | 1.01 | 0.88, 1.17 | 0.8 | - | - | - | - | - | - | 0.87 | 0.74, 1.03 | 0.11 |
| **Neighborhood Vandalism** | 1.02 | 0.92, 1.13 | 0.700 | 1.02 | 0.92, 1.13 | 0.700 | 1.03 | 0.93, 1.15 | 0.5 | 1.01 | 0.91, 1.12 | 0.800 | 1.01 | 0.91, 1.13 | 0.800 | 1.02 | 0.92, 1.14 | 0.7 | 0.99 | 0.87, 1.13 | 0.900 | 0.99 | 0.87, 1.13 | >0.9 | 1.00 | 0.87, 1.14 | >0.9 |
| PGS-AD | - | - | - | 1.13 | 0.95, 1.35 | 0.200 | 1.15 | 0.96, 1.37 | 0.12 | - | - | - | 1.11 | 0.94, 1.33 | 0.200 | 1.12 | 0.94, 1.34 | 0.2 | - | - | - | 1.09 | 0.86, 1.39 | 0.500 | 1.11 | 0.87, 1.41 | 0.4 |
| Neighborhood Vandalism* PGS-AD | - | - | - | - | - | - | 0.93 | 0.82, 1.04 | 0.2 | - | - | - | - | - | - | 0.95 | 0.84, 1.07 | 0.400 | - | - | - | - | - | - | 0.93 | 0.79, 1.09 | 0.4 |
| **Neighborhood Belonging** | 1.08 | 0.97, 1.19 | 0.200 | 1.07 | 0.97, 1.18 | 0.200 | 1.07 | 0.97, 1.19 | 0.2 | 1.08 | 0.97, 1.19 | 0.200 | 1.07 | 0.97, 1.19 | 0.200 | 1.07 | 0.97, 1.19 | 0.200 | 0.97 | 0.84, 1.11 | 0.600 | 0.97 | 0.84, 1.11 | 0.600 | 0.97 | 0.84, 1.12 | 0.7 |
| PGS-AD | - | - | - | 1.13 | 0.95, 1.35 | 0.200 | 1.13 | 0.95, 1.35 | 0.2 | - | - | - | 1.11 | 0.93, 1.33 | 0.200 | 1.11 | 0.93, 1.33 | 0.200 | - | - | - | 1.09 | 0.86, 1.39 | 0.500 | 1.10 | 0.87, 1.40 | 0.4 |
| Neighborhood Belonging * PGS-AD | - | - | - | - | - | - | 0.99 | 0.88, 1.11 | 0.9 | - | - | - | - | - | - | 0.99 | 0.89, 1.12 | >0.9 | - | - | - | - | - | - | 0.90 | 0.77, 1.05 | 0.2 |
| **Neighborhood Cleanness** | 1.02 | 0.91, 1.13 | 0.800 | 1.02 | 0.91, 1.13 | 0.700 | 1.02 | 0.92, 1.14 | 0.7 | 1.01 | 0.90, 1.12 | 0.900 | 1.01 | 0.90, 1.12 | 0.900 | 1.01 | 0.91, 1.13 | 0.8 | 1.01 | 0.88, 1.16 | 0.900 | 1.02 | 0.89, 1.17 | 0.800 | 1.03 | 0.90, 1.18 | 0.7 |
| PGS-AD | - | - | - | 1.13 | 0.95, 1.35 | 0.200 | 1.14 | 0.95, 1.36 | 0.2 | - | - | - | 1.11 | 0.94, 1.33 | 0.200 | 1.12 | 0.94, 1.35 | 0.2 | - | - | - | 1.09 | 0.86, 1.39 | 0.500 | 1.14 | 0.89, 1.46 | 0.3 |
| Neighborhood Cleanness * PGS-AD | - | - | - | - | - | - | 0.98 | 0.86, 1.12 | 0.8 | - | - | - | - | - | - | 0.97 | 0.85, 1.11 | 0.7 | - | - | - | - | - | - | 0.9 | 0.77, 1.05 | 0.2 |
| **Neighborhood Vacant** | 0.95 | 0.85, 1.06 | 0.400 | 0.95 | 0.86, 1.06 | 0.400 | 0.96 | 0.86, 1.08 | 0.500 | 0.95 | 0.86, 1.06 | 0.400 | 0.96 | 0.86, 1.07 | 0.500 | 0.97 | 0.87, 1.08 | 0.6 | 0.99 | 0.86, 1.15 | >0.9 | 1.00 | 0.86, 1.15 | >0.9 | 0.99 | 0.85, 1.14 | 0.9 |
| PGS-AD | - | - | - | 1.13 | 0.95, 1.35 | 0.200 | 1.15 | 0.96, 1.37 | 0.130 | - | - | - | 1.11 | 0.93, 1.33 | 0.200 | 1.12 | 0.94, 1.34 | 0.2 | - | - | - | 1.09 | 0.86, 1.39 | 0.500 | 1.07 | 0.84, 1.36 | 0.6 |
| Neighborhood Vacant * PGS-AD | - | - | - | - | - | - | 0.94 | 0.83, 1.06 | 0.300 | - | - | - | - | - | - | 0.96 | 0.85, 1.08 | 0.5 | - | - | - | - | - | - | 1.12 | 0.95, 1.32 | 0.2 |

All Models adjusted for Age, Sex, Education, Poverty Status, APOE E4 Status, Social Ladder, and Baseline Wave.

**Supplemental Table 6.** Hazard Ratios from survival analysis comparing binary Neighborhood disadvantage index with additional covariates, stratified by Ancestry: risk of incident cognitive impairment (CIND and dementia), CIND and dementia in the US Health and Retirement Study (2008-2010 Waves).

| | **Cognitive Impairment vs. Normal Cognition, European Ancestry (n=6,035)** | | | | | | | | | **CIND vs. Normal Cognition, European Ancestry (n=5,966)** | | | | | | | | | **Dementia vs. Non-dementia, European Ancestry (n=6,689)** | | | | | | | | |
|---|---|---|---|---|---|---|---|---|---|---|---|---|---|---|---|---|---|---|---|---|---|---|---|---|---|---|---|
| | **Model 1** | | | **Model 2** | | | **Model 3** | | | **Model 1** | | | **Model 2** | | | **Model 3** | | | **Model 1** | | | **Model 2** | | | **Model 3** | | |
| | **HR** | **95% CI** | **p-value** | **HR** | **95% CI** | **p-value** | **HR** | **95% CI** | **p-value** | **HR** | **95% CI** | **p-value** | **HR** | **95% CI** | **p-value** | **HR** | **95% CI** | **p-value** | **HR** | **95% CI** | **p-value** | **HR** | **95% CI** | **p-value** | **HR** | **95% CI** | **p-value** |
| **Neighborhood disadvantage index** | | | | | | | | | | | | | | | | | | | | | | | | | | | |
| The least disadvantaged neighborhoods (<=0) | Ref | Ref | Ref | Ref | Ref | Ref | Ref | Ref | Ref | Ref | Ref | Ref | Ref | Ref | Ref | Ref | Ref | Ref | Ref | Ref | Ref | Ref | Ref | Ref | Ref | Ref | Ref |
| The most disadvantaged neighborhoods (>0) | 1.14 | 1.03, 1.27 | **0.012** | 1.15 | 1.03, 1.27 | 0.010 | 1.16 | 1.03, 1.30 | **0.014** | 1.13 | 1.02, 1.26 | **0.019** | 1.14 | 1.02, 1.26 | 0.017 | 1.15 | 1.02, 1.30 | **0.022** | 1.29 | 1.05, 1.57 | **0.014** | 1.28 | 1.05, 1.57 | **0.015** | 1.28 | 1.01, 1.61 | **0.039** |
| **Age** | 1.08 | 1.07, 1.09 | **<0.001** | 1.08 | 1.07, 1.09 | **<0.001** | 1.08 | 1.07, 1.09 | **<0.001** | 1.08 | 1.07, 1.08 | **<0.001** | 1.08 | 1.07, 1.09 | **<0.001** | 1.08 | 1.07, 1.09 | **<0.001** | 1.11 | 1.10, 1.13 | **<0.001** | 1.11 | 1.10, 1.13 | **<0.001** | 1.11 | 1.10, 1.13 | **<0.001** |
| **Sex** | | | | | | | | | | | | | | | | | | | | | | | | | | | |
| Female | Ref | Ref | Ref | Ref | Ref | Ref | Ref | Ref | Ref | Ref | Ref | Ref | Ref | Ref | Ref | Ref | Ref | Ref | Ref | Ref | Ref | Ref | Ref | Ref | Ref | Ref | Ref |
| Male | 1.30 | 1.17, 1.44 | **<0.001** | 1.29 | 1.17, 1.43 | **<0.001** | 1.29 | 1.17, 1.43 | **<0.001** | 1.30 | 1.17, 1.44 | **<0.001** | 1.30 | 1.17, 1.44 | **<0.001** | 1.30 | 1.17, 1.44 | **<0.001** | 1.15 | 0.94, 1.41 | 0.200 | 1.16 | 0.94, 1.42 | 0.200 | 1.16 | 0.94, 1.42 | 0.200 |
| **Education** | | | | | | | | | | | | | | | | | | | | | | | | | | | |
| Above High School/GED | Ref | Ref | Ref | Ref | Ref | Ref | Ref | Ref | Ref | Ref | Ref | Ref | Ref | Ref | Ref | Ref | Ref | Ref | Ref | Ref | Ref | Ref | Ref | Ref | Ref | Ref | Ref |
| High School/GED | 1.59 | 1.41, 1.79 | **<0.001** | 1.59 | 1.42, 1.79 | **<0.001** | 1.59 | 1.42, 1.79 | **<0.001** | 1.62 | 1.43, 1.82 | **<0.001** | 1.62 | 1.44, 1.83 | **<0.001** | 1.62 | 1.44, 1.83 | **<0.001** | 1.64 | 1.26, 2.12 | **<0.001** | 1.65 | 1.27, 2.14 | **<0.001** | 1.65 | 1.27, 2.14 | **<0.001** |
| Less than High School/GED | 2.84 | 2.40, 3.37 | **<0.001** | 2.85 | 2.40, 3.37 | **<0.001** | 2.84 | 2.40, 3.37 | **<0.001** | 2.90 | 2.44, 3.45 | **<0.001** | 2.91 | 2.44, 3.46 | **<0.001** | 2.91 | 2.44, 3.46 | **<0.001** | 4.06 | 2.99, 5.52 | **<0.001** | 4.10 | 3.02, 5.58 | **<0.001** | 4.11 | 3.02, 5.58 | **<0.001** |
| **Poverty Status (Below)** | | | | | | | | | | | | | | | | | | | | | | | | | | | |
| Above Poverty threshold | Ref | Ref | Ref | Ref | Ref | Ref | Ref | Ref | Ref | Ref | Ref | Ref | Ref | Ref | Ref | Ref | Ref | Ref | Ref | Ref | Ref | Ref | Ref | Ref | Ref | Ref | Ref |
| Below Poverty threshold | 1.16 | 0.89, 1.50 | 0.300 | 1.15 | 0.89, 1.49 | 0.300 | 1.15 | 0.89, 1.49 | 0.300 | 1.17 | 0.90, 1.52 | 0.200 | 1.17 | 0.90, 1.52 | 0.300 | 1.17 | 0.90, 1.52 | 0.300 | 1.84 | 1.24, 2.74 | **0.003** | 1.89 | 1.27, 2.81 | **0.002** | 1.89 | 1.27, 2.81 | **0.002** |
| **APOE E4 status (No copy)** | | | | | | | | | | | | | | | | | | | | | | | | | | | |
| No copies of e4 | Ref | Ref | Ref | Ref | Ref | Ref | Ref | Ref | Ref | Ref | Ref | Ref | Ref | Ref | Ref | Ref | Ref | Ref | Ref | Ref | Ref | Ref | Ref | Ref | Ref | Ref | Ref |
| Any copies of e4 | 1.44 | 1.30, 1.60 | **<0.001** | 1.43 | 1.29, 1.59 | **<0.001** | 1.43 | 1.29, 1.59 | **<0.001** | 1.45 | 1.30, 1.61 | **<0.001** | 1.44 | 1.29, 1.60 | **<0.001** | 1.44 | 1.29, 1.60 | **<0.001** | 2.07 | 1.70, 2.52 | **<0.001** | 2.06 | 1.70, 2.51 | **<0.001** | 2.06 | 1.70, 2.51 | **<0.001** |
| **Social Ladder** | 0.95 | 0.92, 0.98 | **0.001** | 0.95 | 0.92, 0.98 | **0.001** | 0.95 | 0.92, 0.98 | **0.001** | 0.95 | 0.92, 0.98 | **0.001** | 0.95 | 0.92, 0.98 | **0.001** | 0.95 | 0.92, 0.98 | **0.001** | 1.00 | 0.94, 1.07 | >0.9 | 1.01 | 0.95, 1.07 | 0.900 | 1.01 | 0.95, 1.07 | 0.900 |
| **Baseline wave** | | | | | | | | | | | | | | | | | | | | | | | | | | | |
| Wave 1 (2008) | Ref | Ref | Ref | Ref | Ref | Ref | Ref | Ref | Ref | Ref | Ref | Ref | Ref | Ref | Ref | Ref | Ref | Ref | Ref | Ref | Ref | Ref | Ref | Ref | Ref | Ref | Ref |
| Wave 2 (2010) | 0.86 | 0.78, 0.94 | **0.002** | 0.86 | 0.78, 0.95 | **0.002** | 0.86 | 0.78, 0.95 | **0.002** | 0.86 | 0.78, 0.95 | **0.002** | 0.86 | 0.78, 0.95 | **0.003** | 0.86 | 0.78, 0.95 | **0.003** | 0.81 | 0.67, 0.97 | **0.026** | 0.81 | 0.67, 0.97 | **0.025** | 0.81 | 0.67, 0.97 | **0.026** |
| **PGS-AD** | | | | | | | | | | | | | | | | | | | | | | | | | | | |
| Below 75% | - | - | - | Ref | Ref | Ref | Ref | Ref | Ref | - | - | - | Ref | Ref | Ref | Ref | Ref | Ref | - | - | - | Ref | Ref | Ref | Ref | Ref | Ref |
| Above 75% | - | - | - | 1.14 | 1.02, 1.27 | **0.021** | 1.16 | 1.01, 1.32 | **0.032** | - | - | - | 1.13 | 1.01, 1.27 | 0.029 | 1.15 | 1.01, 1.32 | **0.042** | - | - | - | 1.17 | 0.94, 1.46 | 0.150 | 1.16 | 0.89, 1.52 | 0.300 |
| **Neighborhood* PGS-AD** | - | - | - | - | - | - | 0.95 | 0.75, 1.21 | 0.7 | - | - | - | - | - | - | 0.95 | 0.75, 1.21 | 0.7 | - | - | - | - | - | - | 1.02 | 0.65, 1.60 | >0.9 |
| RERI: The most disadvantaged neighborhoods*PGS-AD Above 75% | - | - | - | - | - | - | -0.04 | -0.32, 0.25 | | - | - | - | - | - | - | -0.04 | -0.33, 0.25 | | - | - | - | - | - | - | 0.08 | -0.53, 0.68 | |
| **Smoking status** | | | | | | | | | | | | | | | | | | | | | | | | | | | |
| Never Smoker | Ref | Ref | Ref | Ref | Ref | Ref | Ref | Ref | Ref | Ref | Ref | Ref | Ref | Ref | Ref | Ref | Ref | Ref | Ref | Ref | Ref | Ref | Ref | Ref | Ref | Ref | Ref |
| Current Smoker | 1.28 | 1.08, 1.52 | **0.004** | 1.29 | 1.08, 1.53 | **0.004** | 1.29 | 1.08, 1.53 | **0.004** | 1.29 | 1.08, 1.54 | **0.004** | 1.29 | 1.09, 1.54 | **0.004** | 1.29 | 1.09, 1.54 | **0.004** | 1.06 | 0.72, 1.55 | 0.800 | 1.04 | 0.71, 1.53 | 0.800 | 1.04 | 0.71, 1.53 | 0.800 |
| Former Smoke | 1.07 | 0.97, 1.19 | 0.200 | 1.07 | 0.96, 1.19 | 0.200 | 1.07 | 0.96, 1.18 | 0.200 | 1.07 | 0.96, 1.19 | 0.200 | 1.07 | 0.96, 1.18 | 0.200 | 1.07 | 0.96, 1.18 | 0.200 | 1.02 | 0.83, 1.25 | 0.900 | 1.01 | 0.83, 1.24 | >0.9 | 1.01 | 0.83, 1.24 | >0.9 |
| **Drinking (# drinks/day when drinks)** | 1.01 | 0.97, 1.05 | 0.800 | 1.01 | 0.97, 1.05 | 0.700 | 1.01 | 0.97, 1.05 | 0.700 | 1.01 | 0.97, 1.05 | 0.700 | 1.01 | 0.97, 1.05 | 0.700 | 1.01 | 0.97, 1.05 | 0.600 | 1.01 | 0.92, 1.11 | 0.800 | 1.01 | 0.92, 1.11 | 0.800 | 1.01 | 0.92, 1.11 | 0.800 |
| **Depression** | 1.09 | 1.06, 1.12 | **<0.001** | 1.09 | 1.06, 1.12 | **<0.001** | 1.09 | 1.06, 1.12 | <0.001 | 1.09 | 1.06, 1.12 | **<0.001** | 1.09 | 1.06, 1.12 | **<0.001** | 1.09 | 1.06, 1.12 | <0.001 | 1.11 | 1.05, 1.17 | **<0.001** | 1.11 | 1.06, 1.17 | **<0.001** | 1.11 | 1.06, 1.17 | <0.001 |
| **BMI** | 0.99 | 0.98, 1.00 | 0.200 | 1.00 | 0.99, 1.00 | 0.300 | 1.00 | 0.99, 1.00 | 0.3 | 1.00 | 0.99, 1.00 | 0.300 | 1.00 | 0.99, 1.01 | 0.400 | 1.00 | 0.99, 1.01 | 0.4 | 0.97 | 0.95, 0.99 | **0.005** | 0.97 | 0.95, 0.99 | **0.006** | 0.97 | 0.95, 0.99 | **0.006** |
| **Ever have Diabetes** | | | | | | | | | | | | | | | | | | | | | | | | | | | |
| No | Ref | Ref | Ref | Ref | Ref | Ref | Ref | Ref | Ref | Ref | Ref | Ref | Ref | Ref | Ref | Ref | Ref | Ref | Ref | Ref | Ref | Ref | Ref | Ref | Ref | Ref | Ref |
| Yes | 1.14 | 1.0, 1.30 | 0.060 | 1.12 | 0.98, 1.28 | 0.092 | 1.12 | 0.98, 1.28 | 0.092 | 1.16 | 1.01, 1.33 | **0.030** | 1.15 | 1.00, 1.31 | **0.050** | 1.15 | 1.00, 1.31 | 0.050 | 1.06 | 0.80, 1.39 | 0.700 | 1.05 | 0.80, 1.38 | 0.700 | 1.05 | 0.80, 1.38 | 0.700 |
| **Chronic Condition** | | | | | | | | | | | | | | | | | | | | | | | | | | | |
| None | Ref | Ref | Ref | Ref | Ref | Ref | Ref | Ref | Ref | Ref | Ref | Ref | Ref | Ref | Ref | Ref | Ref | Ref | Ref | Ref | Ref | Ref | Ref | Ref | Ref | Ref | Ref |
| 1 - 2 | 0.99 | 0.84, 1.17 | >0.9 | 1.00 | 0.84, 1.18 | >0.9 | 1.00 | 0.84, 1.18 | >0.9 | 1.00 | 0.84, 1.19 | >0.9 | 1.01 | 0.85, 1.19 | >0.9 | 1.01 | 0.85, 1.19 | >0.9 | 0.78 | 0.56, 1.07 | 0.130 | 0.78 | 0.57, 1.08 | 0.130 | 0.78 | 0.57, 1.08 | 0.130 |
| >= 3 | 1.16 | 0.96, 1.39 | 0.120 | 1.17 | 0.98, 1.41 | 0.090 | 1.17 | 0.98, 1.41 | 0.088 | 1.16 | 0.96, 1.40 | 0.130 | 1.17 | 0.97, 1.41 | 0.100 | 1.17 | 0.97, 1.41 | 0.100 | 0.82 | 0.58, 1.17 | 0.300 | 0.82 | 0.58, 1.17 | 0.300 | 0.82 | 0.58, 1.17 | 0.300 |
| | **Cognitive Impairment vs. Normal Cognition, African Ancestry (n=696)** | | | | | | | | | **CIND vs. Normal Cognition, African Ancestry (n=688)** | | | | | | | | | **Dementia vs. Non-dementia, African Ancestry (n=960)** | | | | | | | | |
| | **Model 1** | | | **Model 2** | | | **Model 3** | | | **Model 1** | | | **Model 2** | | | **Model 3** | | | **Model 1** | | | **Model 2** | | | **Model 3** | | |
| | **HR** | **95% CI** | **p-value** | **HR** | **95% CI** | **p-value** | **HR** | **95% CI** | **p-value** | **HR** | **95% CI** | **p-value** | **HR** | **95% CI** | **p-value** | **HR** | **95% CI** | **p-value** | **HR** | **95% CI** | **p-value** | **HR** | **95% CI** | **p-value** | **HR** | **95% CI** | **p-value** |
| **Neighborhood disadvantage index** | | | | | | | | | | | | | | | | | | | | | | | | | | | |
| The least disadvantaged neighborhoods (<=0) | Ref | Ref | Ref | Ref | Ref | Ref | Ref | Ref | Ref | Ref | Ref | Ref | Ref | Ref | Ref | Ref | Ref | Ref | Ref | Ref | Ref | Ref | Ref | Ref | Ref | Ref | Ref |

| | | | | | | | | | | | | | | | | | | | | | | | | | | | |
|---|---|---|---|---|---|---|---|---|---|---|---|---|---|---|---|---|---|---|---|---|---|---|---|---|---|---|---|
| The most disadvantaged neighborhoods (>0) | 1.04 | 0.82, 1.33 | 0.700 | 1.02 | 0.80, 1.29 | 0.900 | 1.13 | 0.85, 1.51 | 0.400 | 1.04 | 0.82, 1.32 | 0.800 | 1.01 | 0.79, 1.29 | >0.9 | 1.11 | 0.83, 1.47 | 0.500 | 1.29 | 0.93, 1.78 | 0.130 | 1.29 | 0.93, 1.79 | 0.130 | 1.34 | 0.92, 1.96 | 0.120 |
| **Age** | 1.05 | 1.04, 1.07 | **<0.001** | 1.06 | 1.04, 1.07 | **<0.001** | 1.06 | 1.04, 1.07 | **<0.001** | 1.06 | 1.04, 1.07 | **<0.001** | 1.06 | 1.04, 1.07 | **<0.001** | 1.06 | 1.04, 1.07 | **<0.001** | 1.09 | 1.07, 1.11 | **<0.001** | 1.09 | 1.07, 1.12 | **<0.001** | 1.09 | 1.07, 1.12 | **<0.001** |
| **Sex** | | | | | | | | | | | | | | | | | | | | | | | | | | | |
| Female | Ref | Ref | Ref | Ref | Ref | Ref | Ref | Ref | Ref | Ref | Ref | Ref | Ref | Ref | Ref | Ref | Ref | Ref | Ref | Ref | Ref | Ref | Ref | Ref | Ref | Ref | Ref |
| Male | 1.18 | 0.91, 1.52 | 0.200 | 1.17 | 0.90, 1.52 | 0.200 | 1.18 | 0.91, 1.53 | 0.200 | 1.18 | 0.91, 1.53 | 0.200 | 1.18 | 0.91, 1.53 | 0.200 | 1.18 | 0.91, 1.53 | 0.200 | 1.25 | 0.88, 1.75 | 0.200 | 1.28 | 0.91, 1.80 | 0.200 | 1.28 | 0.91, 1.80 | 0.200 |
| **Education** | | | | | | | | | | | | | | | | | | | | | | | | | | | |
| Above High School/GED | Ref | Ref | Ref | Ref | Ref | Ref | Ref | Ref | Ref | Ref | Ref | Ref | Ref | Ref | Ref | Ref | Ref | Ref | Ref | Ref | Ref | Ref | Ref | Ref | Ref | Ref | Ref |
| High School/GED | 1.49 | 1.10, 2.02 | **0.010** | 1.47 | 1.08, 2.00 | **0.014** | 1.46 | 1.07, 1.98 | **0.016** | 1.52 | 1.11, 2.07 | **0.009** | 1.49 | 1.09, 2.04 | **0.013** | 1.48 | 1.08, 2.03 | **0.014** | 2.09 | 1.13, 3.87 | **0.020** | 2.17 | 1.17, 4.03 | **0.014** | 2.15 | 1.16, 4.00 | **0.015** |
| Less than High School/GED | 3.01 | 2.11, 4.31 | **<0.001** | 2.90 | 2.03, 4.15 | **<0.001** | 2.93 | 2.04, 4.19 | **<0.001** | 3.12 | 2.17, 4.48 | **<0.001** | 2.97 | 2.06, 4.27 | **<0.001** | 2.98 | 2.07, 4.29 | **<0.001** | 5.26 | 2.81, 9.83 | **<0.001** | 5.49 | 2.92, 10.3 | **<0.001** | 5.48 | 2.92, 10.3 | **<0.001** |
| **Poverty Status** | | | | | | | | | | | | | | | | | | | | | | | | | | | |
| Above Poverty threshold | Ref | Ref | Ref | Ref | Ref | Ref | Ref | Ref | Ref | Ref | Ref | Ref | Ref | Ref | Ref | Ref | Ref | Ref | Ref | Ref | Ref | Ref | Ref | Ref | Ref | Ref | Ref |
| Below Poverty threshold | 1.50 | 1.12, 2.02 | **0.007** | 1.51 | 1.12, 2.03 | **0.007** | 1.51 | 1.12, 2.04 | **0.007** | 1.49 | 1.11, 2.01 | **0.009** | 1.49 | 1.10, 2.01 | **0.010** | 1.50 | 1.11, 2.02 | **0.009** | 1.57 | 1.08, 2.29 | **0.019** | 1.54 | 1.05, 2.24 | **0.026** | 1.53 | 1.05, 2.24 | **0.027** |
| **APOE E4 status** | | | | | | | | | | | | | | | | | | | | | | | | | | | |
| No copies of e4 | Ref | Ref | Ref | Ref | Ref | Ref | Ref | Ref | Ref | Ref | Ref | Ref | Ref | Ref | Ref | Ref | Ref | Ref | Ref | Ref | Ref | Ref | Ref | Ref | Ref | Ref | Ref |
| Any copies of e4 | 0.94 | 0.74, 1.19 | 0.600 | 0.93 | 0.73, 1.19 | 0.600 | 0.93 | 0.73, 1.18 | 0.600 | 0.92 | 0.72, 1.17 | 0.500 | 0.92 | 0.72, 1.17 | 0.500 | 0.91 | 0.72, 1.17 | 0.500 | 1.37 | 0.99, 1.88 | 0.055 | 1.38 | 1.00, 1.90 | **0.048** | 1.38 | 1.00, 1.90 | **0.051** |
| **Social Ladder** | 0.98 | 0.92, 1.05 | 0.600 | 0.97 | 0.91, 1.04 | 0.400 | 0.97 | 0.91, 1.04 | 0.400 | 0.99 | 0.92, 1.06 | 0.700 | 0.98 | 0.91, 1.05 | 0.500 | 0.98 | 0.92, 1.05 | 0.600 | 1.08 | 0.99, 1.18 | 0.067 | 1.08 | 0.99, 1.18 | 0.075 | 1.08 | 0.99, 1.18 | 0.071 |
| **Baseline wave** | | | | | | | | | | | | | | | | | | | | | | | | | | | |
| Wave 1 (2008) | Ref | Ref | Ref | Ref | Ref | Ref | Ref | Ref | Ref | Ref | Ref | Ref | Ref | Ref | Ref | Ref | Ref | Ref | Ref | Ref | Ref | Ref | Ref | Ref | Ref | Ref | Ref |
| Wave 2 (2010) | 1.17 | 0.92, 1.49 | 0.200 | 1.150 | 0.90, 1.46 | 0.300 | 1.13 | 0.89, 1.45 | 0.3 | 1.17 | 0.92, 1.50 | 0.200 | 1.150 | 0.90, 1.48 | 0.300 | 1.14 | 0.89, 1.46 | 0.3 | 1.05 | 0.77, 1.44 | 0.800 | 1.040 | 0.75, 1.43 | 0.800 | 1.04 | 0.75, 1.43 | 0.800 |
| **PGS-AD** | | | | | | | 1.11 | 0.94, 1.31 | 0.2 | | | | | | | 1.11 | 0.94, 1.31 | 0.2 | | | | | | | | | |
| Below 75% | - | - | - | Ref | Ref | Ref | Ref | Ref | Ref | - | - | - | Ref | Ref | Ref | Ref | Ref | Ref | - | - | - | Ref | Ref | Ref | Ref | Ref | Ref |
| Above 75% | - | - | - | 1.14 | 0.85, 1.52 | 0.400 | 1.45 | 0.94, 2.22 | 0.089 | - | - | - | 1.1 | 0.82, 1.47 | 0.500 | 1.35 | 0.87, 2.09 | 0.200 | - | - | - | 1.03 | 0.68, 1.56 | 0.900 | 1.14 | 0.62, 2.09 | 0.700 |
| **Neighborhood* PGS-AD** | - | - | - | - | - | - | 0.68 | 0.41, 1.14 | 0.150 | - | - | - | - | - | - | 0.72 | 0.43, 1.23 | 0.200 | - | - | - | - | - | - | 0.84 | 0.39, 1.79 | 0.600 |
| RERI: The most disadvantaged neighborhoods*PGS-AD Above 75% | - | - | - | - | - | - | -0.46 | -1.16, 0.23 | | - | - | - | - | - | - | -0.38 | -1.04, 0.29 | | - | - | - | - | - | - | -0.20 | -1.13, 0.73 | |
| **Smoking status** | | | | | | | | | | | | | | | | | | | | | | | | | | | |
| Never Smoker | Ref | Ref | Ref | Ref | Ref | Ref | Ref | Ref | Ref | Ref | Ref | Ref | Ref | Ref | Ref | Ref | Ref | Ref | Ref | Ref | Ref | Ref | Ref | Ref | Ref | Ref | Ref |
| Current Smoker | 1.23 | 0.88, 1.72 | 0.200 | 1.27 | 0.91, 1.78 | 0.200 | 1.25 | 0.89, 1.75 | 0.200 | 1.25 | 0.89, 1.76 | 0.200 | 1.29 | 0.92, 1.81 | 0.140 | 1.28 | 0.91, 1.79 | 0.200 | 1.42 | 0.85, 2.37 | 0.200 | 1.44 | 0.86, 2.42 | 0.200 | 1.45 | 0.86, 2.43 | 0.200 |
| Former Smoke | 0.95 | 0.73, 1.24 | 0.700 | 1.00 | 0.76, 1.30 | >0.9 | 0.98 | 0.75, 1.28 | 0.900 | 0.92 | 0.71, 1.20 | 0.500 | 0.96 | 0.73, 1.25 | 0.700 | 0.94 | 0.72, 1.24 | 0.700 | 1.44 | 1.01, 2.05 | **0.041** | 1.46 | 1.02, 2.09 | 0.037 | 1.45 | 1.02, 2.08 | **0.040** |
| **Drinking (# drinks/day when drinks)** | 1.03 | 0.94, 1.12 | 0.600 | 1.03 | 0.94, 1.12 | 0.600 | 1.02 | 0.94, 1.12 | 0.600 | 1.03 | 0.94, 1.13 | 0.500 | 1.03 | 0.94, 1.13 | 0.500 | 1.03 | 0.94, 1.13 | 0.500 | 0.99 | 0.86, 1.13 | 0.900 | 0.98 | 0.86, 1.13 | 0.800 | 0.98 | 0.86, 1.13 | 0.800 |
| **Depression** | 1.07 | 1.01, 1.14 | **0.019** | 1.08 | 1.01, 1.14 | **0.016** | 1.08 | 1.01, 1.14 | 0.017 | 1.08 | 1.01, 1.14 | **0.016** | 1.08 | 1.02, 1.15 | **0.014** | 1.08 | 1.01, 1.14 | 0.015 | 1.14 | 1.06, 1.23 | **<0.001** | 1.15 | 1.07, 1.24 | **<0.001** | 1.15 | 1.07, 1.24 | <0.001 |
| **BMI** | 0.99 | 0.97, 1.01 | 0.400 | 0.99 | 0.97, 1.01 | 0.400 | 0.99 | 0.97, 1.01 | 0.400 | 0.99 | 0.97, 1.01 | 0.500 | 0.99 | 0.97, 1.01 | 0.400 | 0.99 | 0.97, 1.01 | 0.500 | 0.99 | 0.96, 1.01 | 0.400 | 0.99 | 0.96, 1.01 | 0.400 | 0.99 | 0.96, 1.01 | 0.400 |
| **Ever have Diabetes** | | | | | | | | | | | | | | | | | | | | | | | | | | | |
| No | Ref | Ref | Ref | Ref | Ref | Ref | Ref | Ref | Ref | Ref | Ref | Ref | Ref | Ref | Ref | Ref | Ref | Ref | Ref | Ref | Ref | Ref | Ref | Ref | Ref | Ref | Ref |
| Yes | 1.13 | 0.84, 1.52 | 0.400 | 1.16 | 0.86, 1.57 | 0.300 | 1.16 | 0.86, 1.56 | 0.300 | 1.13 | 0.84, 1.53 | 0.400 | 1.17 | 0.86, 1.58 | 0.300 | 1.16 | 0.85, 1.57 | 0.300 | 0.95 | 0.64, 1.42 | 0.800 | 1.01 | 0.67, 1.50 | >0.9 | 1.01 | 0.67, 1.50 | >0.9 |
| **Chronic Condition** | | | | | | | | | | | | | | | | | | | | | | | | | | | |
| None | Ref | Ref | Ref | Ref | Ref | Ref | Ref | Ref | Ref | Ref | Ref | Ref | Ref | Ref | Ref | Ref | Ref | Ref | Ref | Ref | Ref | Ref | Ref | Ref | Ref | Ref | Ref |
| 1 - 2 | 0.70 | 0.49, 1.01 | 0.058 | 0.72 | 0.49, 1.04 | 0.077 | 0.72 | 0.50, 1.04 | 0.081 | 0.68 | 0.47, 0.99 | **0.043** | 0.69 | 0.48, 1.01 | 0.056 | 0.70 | 0.48, 1.01 | 0.059 | 0.84 | 0.49, 1.44 | 0.500 | 0.85 | 0.49, 1.45 | 0.500 | 0.84 | 0.49, 1.45 | 0.500 |
| >= 3 | 0.96 | 0.63, 1.47 | 0.900 | 0.96 | 0.62, 1.48 | 0.900 | 0.98 | 0.64, 1.51 | >0.9 | 0.96 | 0.62, 1.48 | 0.800 | 0.96 | 0.62, 1.49 | 0.900 | 0.98 | 0.63, 1.52 | >0.9 | 0.82 | 0.45, 1.49 | 0.500 | 0.80 | 0.43, 1.46 | 0.500 | 0.80 | 0.44, 1.48 | 0.500 |
